# Towards the development of a management protocol for Subjective Cognitive Decline: insights from a cross-sectional and longitudinal analyses of multimodal data from a memory clinic

**DOI:** 10.1101/2024.10.15.24315516

**Authors:** Salvatore Mazzeo, Michael Lassi, Sonia Padiglioni, Alberto Arturo Vergani, Valentina Moschini, Maenia Scarpino, Giulia Giacomucci, Rachele Burali, Carmen Morinelli, Carlo Fabbiani, Giulia Galdo, Lorenzo Gaetano Amato, Silvia Bagnoli, Filippo Emiliani, Assunta Ingannato, Benedetta Nacmias, Sandro Sorbi, Antonello Grippo, Alberto Mazzoni, Valentina Bessi

## Abstract

**Introduction:** Subjective cognitive decline (SCD) is considered as an early symptomatic stage of Alzheimer’s disease (AD). This study aimed to develop a management protocol for SCD based on cross-sectional and longitudinal analyses.

**Methods:** We enrolled 440 SCD patients who underwent neurological and neuropsychological assessments, MRI scans, *APOE* genotyping, and AD biomarker evaluations. Patients were followed for a median of 10 years.

**Results:** A link was observed between SCD severity, insomnia, and benzodiazepine use. Aβ pathology was identified in 35% of cases. During follow-up, 87 patients progressed to mild cognitive impairment or dementia. The multi-modal clinical profile of each patient was processed by a machine learning model, predicting progression with 74% accuracy and providing a hierarchy of importance of features predicting progression.

**Discussion:** Our findings support the development of a personalized clinical management protocol for SCD that considers demographic, clinical, cognitive and biological factors for both baseline characterization and prognostic purposes.

## 1. Background

“Losing memory” is a commonly reported experience in elderly [1] and in middle-aged people [2]. Approximately one-third of older adults complain of memory or other cognitive difficulties [3]. This condition has been referred to by various names (subjective cognitive impairment, subjective memory decline, subjective cognitive decline, etc. [4]). The lack of a common terminology and criteria led to contrasting results among studies on individuals complaining memory loss [5–7].

In 2014, the Subjective Cognitive Decline (SCD)-Initiative (SCD-I) defined SCD as a self-experienced persistent decline in cognitive capacity in comparison with the individual’s previously normal status, during which the individual has normal age-, sex-, and education-adjusted performance on standardized cognitive tests [8]. The definition of SCD as a nosological entity allowed researchers to work on the same concept, applying common criteria for study design. As a result, most of the previously conflicting results have been clarified. In particular, a growing body of evidence now supports the higher risk of dementia in SCD patients compared to individuals of the same age without SCD [9]. Recognizing this association, the National Institute of Aging-Alzheimer’s Association (NIA-AA) has included SCD as the first manifestation of symptomatic AD stages, preceding mild cognitive impairment (MCI) [10].

However, not all the patients reporting SCD will develop dementia. Most of them have been defined as ‘‘worried well’’ and do not deteriorate more rapidly than usual in normal aging [11]. In other cases, subjective decline in cognition has been related to conditions such as personality traits, psychiatric conditions, medical disorders, or use of medication [12]. As a result, individuals with SCD constitute a heterogeneous group. Therefore, understanding if SCD is a valid concern is crucial both for people experiencing SCD, both for targeting dementia prevention [13], particularly in the upcoming era of disease modifying therapies (DMTs) for AD [14]. Indeed, it is a shared understanding that DMTs should be administered at the earliest stages of the disease [15]. Consequently, individuals with SCD may represent a target population for DMT to preserve cognitive function [16]. Therefore, there is an urgent need to select cost-effective and easily accessible tools for identifying patients at the early stages of AD, while minimizing the inclusion of patients who will not benefit of treatment for AD.

The relevance of SCD is not limited to early prediction of AD. Independently from progression to objective cognitive impairment, SCD has been related to a variety of poor health outcomes including lower quality of life [17], frailty [18], chronic health conditions [19], and sleep disorders [20]. Nevertheless, despite the high frequency of individuals seeking attention for SCD and the impact of this condition on well-being, there is no consensus on how to manage these patients. This lack of standardization results in significant heterogeneity across different centers, leading to highly variable treatments for each patient. The absence of clear guidelines on which aspects to investigate at baseline can result in the neglect of potentially treatable causes or the underestimation of factors that may indicate a risk of cognitive decline progression [21]. Furthermore, the lack of follow-up protocols may lead to suboptimal intervals between visits: patients at higher risk of progression may not be monitored closely enough, leading to missed opportunities to detect cognitive deterioration; patients at lower risk may be seen too frequently, resulting in unnecessary overcrowding of memory clinics. Here we aimed to address these gaps through a comprehensive description (by a cross-sectional and longitudinal analysis) of a memory clinic cohort of patients with SCD. Specifically, we aimed to:

1. Describe a large cohort of patients with SCD, detailing demographic, clinical, neuropsychological, biological, and personality features, and exploring their interactions.
2. Investigate the proportion of positive AD biomarkers in SCD.
3. Analyze longitudinal data to delineate trajectories of our patient cohort and build a predictive model of progression of cognitive decline.
4. Establish a management protocol for SCD to guide clinical practice in memory clinic settings.

## 2. Methods and Analysis

### 2.1. Patients and assessments

Four-hundred-forty participants have been consecutively recruited among patients referred to the Centre for Alzheimer’s Disease and Adult Cognitive Disorders of Careggi Hospital in Florence between January 1994 and November 2023. Inclusion criteria were: 1) complaining of cognitive decline with a duration of ≥ 6 months; 2) normal functioning on the Activities of Daily Living and the Instrumental Activities of Daily Living scales [22]; 3) unsatisfied criteria for MCI and dementia at baseline [23,24]. Exclusion criteria were: history of head injury, current neurological and/or systemic disease, symptoms of psychosis, major depression, substance use disorder.

All the participants underwent a comprehensive family and clinical history, general and neurological examination and extensive neuropsychological investigation (NPS) (see Supplementary Methods for details of the neuropsychological battery).

Age at onset of symptoms was established as the age at which individuals started experiencing cognitive decline. Age at baseline referred to the age at first neuropsychological examination. Symptom duration was defined as the timeframe between age at onset and age at baseline. Based on SCD-plus criteria proposed by Jessen et al. [8] and confirmed by other works [25,26], we classified patients as “younger” or “older” if they were younger than 60 or 60 years of age or older at onset of symptoms. Other two SCD-plus criteria were collected during neuropsychological examination based on patient interview at first neurological visit: “subjective decline in memory, rather than other domains of cognition” and “concerns (worries) associated with SCD”. Positive family history was defined as one or more first-degree relatives with documented cognitive decline.

Presence or absence of hypertension, dyslipidemia, impaired fasting glucose (IFG), diabetes, current or previous smoking habit, insomnia, use of benzodiazepine, antidepressants (SSRI/SNRI) or anticholinergic drugs were defined based on medical history. Use of anticholinergics was defined according to the AntiCholinergic Burden (ACB) [27] (use of anticholinergics if ACB >1).

Two-hundred-fifty-five patients underwent brain MRI scan on 1.5 Tesla scanners. Presence or absence of white matter hyperintensities as a marker of cerebral small vessel disease (CSVD) was defined according to Fazekas scale [28] (present if Fazekas score ≥ 1).

In a subgroup of 164 patients, we evaluated personality traits and leisure activities. In 196 patients who gave consent for genetic analysis we performed *APOE* genotyping.

A subgroup of 37 patients, recruited between February 2015 and March 2023, underwent CSF AD biomarker (Aβ_42_, Aβ_40_, t-tau, and p-tau) assessment at baseline. Among these, 17 patients also underwent amyloid-PET. Moreover, six patients who refused lumbar puncture only underwent amyloid-PET. In total, 43 patients had an assessment of Aβ pathology.

Patients underwent clinical and neuropsychological follow-up every 12 or 24 months. We defined the progression from SCD to MCI according to the National Institute on Aging-Alzheimer’s Association (NIA-AA) criteria [24] and conversion to AD according to the NIA-AA criteria [23].

We defined reversible or stable SCD as occurring in patients who did not progress to objective cognitive decline or who ceased to report cognitive disturbances during the follow-up period.

All subjects were recruited in accordance with the Declaration of Helsinki and with the ethical standards of the Committee on Human Experimentation of Careggi University Hospital (Florence, Italy). The study was approved by the local Institutional Review Board (“Comitato Etico di Area Vasta Centro” reference 15691oss). All participants in this study provided informed consent, agreeing to participate and to share the results deriving from their data.

### 2.2. Personality traits and cognitive reserve

Personality traits were assessed at baseline using the Big Five Factors Questionnaire that follows a widely accepted five-traits personality model [29]: 1) emotional stability, 2) energy, 3) conscientiousness, 4) agreeableness, and 5) openness to culture and experience.

Premorbid intelligence was estimated using the Short intelligence test (TIB) [30], the Italian equivalent of the National Adult Reading Test [31]. Severity of depressive symptoms was evaluated using the Hamilton Depression Rating Scale (HDRS) [32]. Depressive symptoms were established as present if HDRS was greater than 13. The severity of SCD was investigated using the Memory Assessment Clinics-Questionnaire (MAC-Q) [33].

To quantify involvement in leisure activities, we used a questionnaire based on Yarnold PR et al. [34]. Patients were interviewed to estimate their participation in nine intellectual activities, seven social activities and seven physical activities. The frequency of participation was reported for each activity performed during in each previous decade on a Likert scale ranging from 0 to 5, where 0 refers to never, 1 to occasionally, 2 to monthly, 3 to once a week, 4 to several days per week and 5 to daily. We summed the scores for each activity to generate total scores for intellectual, social, physical, and other activities ranging from 0 to 30.

### 2.3. Blood sample collection and *APOE* genotype analysis

Blood samples was collected by venipuncture into standard polypropylene EDTA test tubes (Sarstedt, Nümbrecht, Germany) at the Neurology Unit of Careggi University Hospital. They were centrifuged within two hours at 1300 rcf at room temperature for 10 minutes, and plasma will be isolated and stored at -80°C until tested at the Laboratory of Neurogenetics at Careggi University Hospital.

A standard automated method (QIAcube, QIAGEN) was used to isolate DNA from peripheral blood samples. *APOE* genotypes were investigated by high-resolution melting analysis [35]. Two sets of PCR primers were designed to amplify the regions encompassing rs7412 [NC_000019.9:g[M13] [GG14]. 45412079C>T] and rs429358 (NC_000019.9:g.45411941T>C). The samples with known *APOE* genotypes, which had been validated by DNA sequencing, were used as standard references.

### 2.4. CSF sample collection and Biomarker analysis

CSF samples were collected at 8:00 a.m. by lumbar puncture at the Neurology Unit of Careggi University Hospital. Samples were immediately centrifuged and stored at -80 °C until performing the analysis at the Laboratory of Neurogenetics of Careggi University Hospital.

Aβ_42_, Aβ_40_, t-tau, and p-tau were measured using a chemiluminescent enzyme immunoassay analyser LUMIPULSE G600 (Fujirebio, Tokyo, Japan). Cut-off values for CSF biomarkers were determined following Fujirebio guidelines (diagnostic sensitivity and specificity using clinical diagnosis and the follow-up golden standard as of November 19^th^, 2018).

### 2.5. Amyloid Pet acquisition and rating

Amyloid PET imaging was performed according to standard national and international guidelines [36], with any of the available fluorine18-labeled tracers (^18^F-Florbetaben [FBB]-Bayer-Pyramal, ^18^F-Flutemetamol [FMM]-General Electric). Images were rated as either positive or negative according to criteria defined by the manufacturers.

### 2.6. Classification of patients according to the ATN classification

Since the biomarker assessment was conducted prior to the most recent revision of the AD criteria [37], we classified our patients according to the ATN framework proposed by the AA in 2018 [38]: patients were rated as A+ if at least one of the amyloid biomarkers (CSF or amyloid PET) revealed the presence of Aβ pathology, and as A– if none of the biomarkers revealed the presence of Aβ pathology. In the case of discordant CSF and Amyloid PET results, we considered only the pathologic result. Patients were classified as T+ or T– if CSF p-tau concentrations were higher or lower than cut-off values, respectively. Patients were classified as N+ if CSF t-tau was higher than cut-off value.

### 2.7. Statistical analysis

#### 2.7.1. Descriptive analyses

Descriptive statistical analyses were conducted using IBM SPSS Statistics Software Version 25 (SPSS Inc., Chicago, USA) and the computing environment R4.2.3 (R Foundation for Statistical Computing, Vienna, 2013). Scores at cognitive tests were reported as z-scores (z-scores were calculated as the raw score of the patient, minus the mean score of Italian general population, divided by the SD of Italian general population). The distributions of variables were assessed using the Shapiro-Wilk test. Patient groups were characterized using medians and interquartile ranges (IQR), frequencies or percentages and 95% confidence intervals (95% C.I.) for non-normally distributed variables and categorical variables, respectively. We employed the non-parametric Mann-Whitney U test for between-group comparisons and Spearman’s correlation coefficient to assess correlations between numeric measures within the groups. Chi-square test was used to compare categorical data. Effect sizes were computed using η^2^ for the Mann-Whitney-U Test, and Cramer’s V for categorical data. We used multiple regression models for multivariate analysis to adjust for potential confounders and to assess the associations between variables. The groups were defined based on dichotomous variables, including: women vs. men, younger vs. older patients, patients with vs. without depressive symptoms, patients with vs. without insomnia, patients who used vs. did not use medications, patients with vs. without worry about SCD, women in menopause vs. women not in menopause, patients with vs. without CSVD, women who used HRT vs. those who did not, patients with vs. without a family history of dementia, and patients with vs. without the *APOE* ε4 allele.

To evaluate the incidence rates of progression to MCI and dementia within our cohort, we calculated the incidence rate for each outcome over the follow-up period. The incidence rate was defined as the number of new cases of MCI that occurred during the follow-up period divided by the total person-years at risk. Person-years at risk were calculated by summing the time each patient remained under observation, starting from their entry into the study until the first occurrence of MCI, loss to follow-up, or the end of the study period, whichever came first. Patients who did not develop MCI contributed their total follow-up time to the person-years calculation. The incidence rate was then calculated using the following formula:

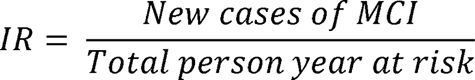

To account for varying follow-up times among patients in our cohort, we calculated the cumulative incidence rates (CIR) of MCI for each year of follow-up. The cumulative incidence rate for each year was determined by dividing the number of new cases of MCI identified during that year by the total number of patients who were still classified as having SCD at the beginning of the year, after subtracting any patients who were lost to follow-up during that period.

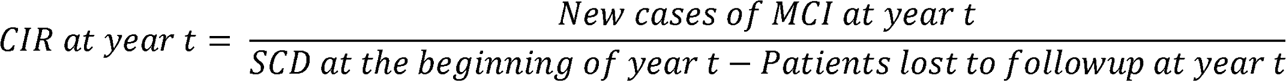

#### 2.7.2. Network analysis

To explore the relationship among features at baseline, we applied a network analysis based on graph theory. To create the networks (based on Markov random field [MRF] modeling), we used the polychoric correlation method for the severity and distress data, using the R-package qgraph [39,40] in combination with Extended Bayesian Information Criterion (EBIC) and the graphical “least absolute shrinkage and selection operator” (glasso) algorithm with a tuning parameter of 0.25 [41]. Missing data were handled pairwise. An undirected graph describes the relations in the MRF, with nodes (rectangles) representing variables and edges (lines) representing interconnection between them. We estimate three centrality indices (betweenness, closeness, strength) and tested for significant differences between edges weights and centrality indices based on 2500 nonparametric bootstrap iterations. Strength indicates which node has the strongest *overall connection,* by measuring the average weight of its edges. Betweenness measures the number of times a node lies on the shortest path between two other nodes. Closeness summarizes the average distance of a node to all other nodes in the network. To estimate the stability of the order of the centrality indices, we used a case-dropping bootstrap technique (based on 2500 bootstrap iterations) together with the correlation stability coefficient (Cs-coefficient). The Cs-coefficient quantifies the maximum proportion of cases or nodes, respectively, can be dropped at random to retain, with 95% certainty, a correlation of at least 0.7 with the centralities of the original network. While no strict cut-of value exists for the Cs-coefficient, its value should be at least 0.25 and preferably higher than 0.5 [42].

#### 2.7.3. Machine learning classification of patient conditions

Demographic, clinical, cognitive, and genetic data were used as input features to a machine learning pipeline, based on the XGboost model [43]. We considered ‘progressing SCD” (pSCD) and ‘stable/reversible SCD’ (r/sSCD) as output labels. The input raw data were subjected to missing data imputation and minority class data oversampling before feeding them to the classifier training. Missing data for continuous variables was imputed using the median value for each feature, while using the most frequent value for categorical/binary variables. Median and mode values of the training set are used to impute the missing values both in the training and testing sets.

The processed features are then submitted to the SMOTE algorithm [44], where the minority class (r/s-SCD in our dataset) is oversampled to match the majority class numerosity by generating artificial samples. The algorithm selects the minority class instance and finds its k nearest minority class neighbors (we set k = 5). It then generates random synthetic instances along the line segments joining these neighbors. By using SMOTE, the effect of class imbalance is hence mitigated, reducing the risks of bias in the trained algorithm. The oversampling is only applied to training samples.

To solve the binary classification problem, the processed training dataset is then submitted to the XGboost algorithm. XGboost is an ensemble learning algorithm, that uses boosting of decision trees to perform the classification. Boosting algorithms sequentially add weak learners (in this case, decision trees) to the ensemble classifier, to improve the performance of the overall model. XGboost implementation of gradient boosted trees has shown state-of-the-art performance in classification problems of tabular data [45].

The training, validation and testing of the model followed a nested cross-validation scheme [46] that consists of two loops of cross-validation. The inner validation loop consisted of a 5-fold cross-validation: at each iteration, 4/5 of the training data are used to train the XGboost model, with different sets of hyperparameters, while 1/5 is used to validate its performance. The model and the hyperparameters yielding the best performances, are then used in the outer validation loop. This loop consists of a leave-one-out cross-validation, performing unbiased testing of the performance of the model on an unseen subject.

The following XGboost hyperparameters were optimized in the cross-validation, using a randomized search approach (with n = 100 iterations): maximum tree depth (possible values: 1,2,3,5,8,10), learning rate (0.01, 0.1, 0.2, 0.3, 0.4), subsample (range between 0.5 and 1, step 0.1), column sample by tree (range between 0.5 and 1, step 0.1), column sample by level (range between 0.5 and 1, step 0.1), and number of estimators (10, 50, 100, 500, 1000).

The performance metric used to optimize hyperparameters was balanced accuracy, while the following performance metrics were computed for the test set performance: balanced accuracy, F1 score, precision, recall, area under the ROC curve (AUC) [47]. Performance metrics were bootstrapped to computed 95% confidence intervals (10e3 iterations).

To inspect how each feature contributed to the output classification on the test set, we computed SHapley Additive exPlanations (SHAP) values [48]. SHAP values are a method used in machine learning to explain the output of a model by assigning importance values to each feature. These values are based on game theory and provide insights into how each feature contributes to the model’s predictions, by giving insights on both local and global explanations of the model’s output. Average absolute SHAP value was computed for each feature, as an indicator of the average feature importance. Confidence intervals were computed by bootstrapping the SHAP values 1000 times.

SHAP values were also visualized per each feature to observe the directionality of the decision (whether a higher value of the considered feature positively or negatively influenced the model output). Feature importance based on SHAP was compared among features, by means of a bootstrap test (1000 bootstrap iterations). P-values were corrected with the Bonferroni correction to account for multiple comparisons. The significance level was set at 0.05.

## 3. Results

### 3.1 Cross-sectional analysis

#### 3.1.1 Characteristics of SCD patients at baseline

Features of 440 patients who underwent at least one baseline visit are summarized in Table 1. Age at onset, age at baseline visit and duration of symptoms were not normally distributed. Median age at onset and at baseline were 59.0 years (IQR = 13.0, min:max = 34:86) and 62.8 years (IQR = 14.0, min:max = 34:84) respectively, with a median latency period from onset to the baseline visit (symptom duration) of 2.8 (IQR = 3.6). The vast majority of patients were women (68.9% vs. 31.1%, one-sample t-test for proportion: p < 0.001). As expected, considering the age at the baseline visit, the majority of women were in menopause (89.2%). Family history of AD was reported by about half of patients (56%). Regarding clinical features, only 25 (6.9%) patients had depressive symptoms. 60% of patients had CSVD at MRI scan. The majority of patients with CSVD (55.1%) had a mild grade (Fazekas 1), only eight and three patients had respectively moderate and severe (Fazekas 2 and 3).

**Table 1.** Demographic features of SCD patients.

|  |  |
| --- | --- |
| **Demographics** |  |
| Age at onset | 59.0 (14.0) |
| Age at onset > 60 | 207 (47.0) |
| Age at first visit | 62.8 (14.0) |
| Symptoms duration | 2.8 (3.6) |
| Women | 303 (68.9) |
| Women in menopause | 231 (89.2) |
| Menopause age | 50.0 (5.0) |
| Family history of dementia | 241 (56.6) |
| **Clinical** |  |
| Women with use of HRT | 71 (31.0) |
| Early menopause | 6 (2.6) |
| Insomnia | 224 (54.6) |
| HDRS | 5.0 (6.0) |
| Depressive symptoms | 25/361 (6.9) |
| MMSE | 27.9 (7.8) |
| MAC-Q | 26.0 (4.0) |
| APOE ε4 | 57 (29.1) |
| Confirmation by caregiver | 50 (63.3) |
| Memory complaint | 196 (92.0) |
| Worry about SCD | 111 (74.0) |
| **Cognitive reserve** |  |
| Years of education | 13.0 (9.0) |
| TIB | 112.8 (6.5) |
| **Drugs** |  |
| BZD use | 58 (14.2) |
| SSRI/SNRI use | 69 (16.8) |
| Anticholinergic use | 94 (22.9) |
| **Cardiovascular features** |  |
| Hypertension | 142 (38.7) |
| Dyslipidemia | 98 (29.3) |
| IFG or diabetes | 33 (10.1) |
| Diabetes | 14 (4.3) |
| Previous or current smoking habit | 161 (42.6) |
| Current smoking habit | 34 (9.0) |
| CSVD (Fazekas > 1) | 135 (60.0) |

Values quoted in table are mean ( ± SD) or percentages. HRT: hormone replacement therapy; HDRS: Hamilton Depressive Rating Scale; MAC-Q: Memory Assessment Clinics-Questionnaire; TIB: *Test di intelligenza breve*; BZD: benzodiazepine; SSRI/SNRI: selective serotonin reuptake inhibitors/serotonin noradrenaline reuptake inhibitors; ACB: anticholinergic burden; IFG: impaired fasting glucose; CSVD: cerebral small vessel disease.

#### 3.1.2 Comparison between women and men

Women were younger at onset (57.5 [14.0] vs. 62.0 [12.7], p = 0.016) and at baseline (62.1 [13.1] vs. 65.6 [13.7], p = 0.022), had lower proportion of *APOE* ε4 (17.7% vs. 40.0%, χ² = 4.4, p = 0.036, V = 0.150), higher MAC-Q (26.0 [3.0] vs. 25.0 [2.0], p = 0.002) and HDRS (27.0 [7.0] vs. 25.0 [5], p = 0.002) and lower scores in TIB (< 0.001). Women also had a higher proportion of insomnia (58.5% vs. 48.1%, χ² = 5.2, p = 0.019, V = 0.116), use of SSRI/SNRI (20.42% vs. 8.7%, χ² = 8.5, p = 0.004, V = 0.144) and anticholinergic drugs (26.3% vs. 15.1%, χ² = 6.2, p = 0.012, V = 0.123). Regarding NPS, after age- and education adjustment, women had lower score in tests assessing language (Token test, p-_adj_ = 0.001), executive function (TMT-B, p-_adj_ = 0.041) and spatial span (p-_adj_ = 0.001).

As expected, regarding cardiovascular features, women had lower proportion of hypertension (32.8% vs. 51.3%, χ² = 11.5, p = 0.001, V = 0.177), lower proportion of IFG (7.5% vs. 15.6%, χ² = 5.4, p = 0.020, V = 0.128), lower proportion of previous smoking habit (35.7% vs. 57.5%, χ² = 15.8, p < 0.001, V = 0.206). To exclude that these differences were due to the higher age of men, we performed a multiple regression analysis that confirmed that hypertension (p-_adj_ = 0.017) and previous smoking habits (p-_adj_ = 0.001) were associated with gender independently from age.

#### 3.1.3 Comparison between younger and older patients

Older patients (age at onset > 60) had less years of education (12.0 [7.0] vs 13.0 [6.0], p < 0.001) and lower premorbid intelligence (112.4 [5.4] vs. 113.3 [8.1], p = 0.019), shorter symptom duration (3.6 [4.7] vs. 2.4 [2.4], p < 0.001) and higher proportion of *APOE* ε4 (36.5 % vs. 23.4 %, χ² = 5.4, p = 0.046, V = 3.973) than younger patients. As expected, CSVD (72.6% vs. 48.7%, χ² = 13.3, p < 0.001, V = 0.244), hypertension (53.2% vs. 26.0%, χ² = 28.5, p < 0.001, V = 0.279), and dyslipidemia (37.2 % vs. 23.0%, χ² = 8.0, p = 0.005, V = 0.155) were higher in older patients.

#### 3.1.4 Comparison between patients with and without depressive symptoms

The group of patients with depressive symptoms (HDRS>13) had a higher proportion of worry about SCD (100% vs. 68.9%, χ² = 6.8, p = 0.009, V = 0.225), a lower proportion of women in menopause (70.6% vs. 92.8%, χ² = 9.56, p = 0.002, V = 0.206), and a higher proportion of use of SSRI/SNRI (34.8% vs. 16.6%, χ² = 4.8, p = 0.029, V = 0.119). Patients with depressive symptoms also had lower scores in MMSE (26.2 [IQR 3.2] vs. 29.5 [IQR 3.8], p = 0.005) and in neuropsychological assessing: spatial short-term memory (spatial span [p = 0.009]), ecological memory (RMBT [p = 0.044]), executive function (TMT-B [p = 0.008], TMT-BA [p = 0.003]), and visuospatial abilities (ROCFC [p = 0.021], figure copy [p = 0.001]). There were no other differences between the groups. In particular, patients with depressive symptoms did not have a higher proportion of BZD compared to patients without (8.7% vs. 14.8%, p = 0.422).

#### 3.1.5 Comparison between patients with and without insomnia

As mentioned above, insomnia was more frequent in women than in men (p = 0.019). Patients with sleep disorder also have a higher prevalence of CSVD (65.8% vs. 52.0%, p = 0.040, V = 0.140) and use of BZD (6.6% vs. 20.6%, χ² = 16.2, p < 0.001, V = 0.201), and a lower proportion of previous smoking habit (37.6% vs. 58.5%, p = 0.035, V = 0.110). In women, sleep disorder was associated with a higher proportion of use of HRT (23.1% vs. 36.8%, χ² = 4.7, p = 0.029, V = 0.145). Patients with sleep disorder had higher score in MAC-Q (26.0 [4.0] vs. 25.0 [2] p = 0.003), HDRS (28.0 [6.0] vs. 25.0 [5.0] p < 0.001) and lower scores in TIB (112.0 [7.7] vs. 113.3 [6.2], p = 0.044) and in a NPS for executive function (TMT-B, p = 0.045) and verbal short-term memory (digit span, p = 0.026).

#### 3.1.6 Associations with use of medications

Patients who were taking BZD had lower scores in emotional stability (41.0 [15.0] vs. 51.0 [15.0], p = 0.020) and in NPS assessing attention (MFTC, p < 0.001) and constructional praxis (ROCFC, p = 0.018). Patients with use of anticholinergics had a higher score in HDRS (27.0 [6.0] vs. 26.0 [5.0], p = 0.029), and lower scores in attention (MFTC, p < 0.001) and constructional praxis (CDT, p = 0.045). There were no differences between patients who were taking SSRI/SNRI and patients who were not.

#### 3.1.7 Other comparisons

Patients with worry about SCD had longer symptom duration (3.5 [5.0] vs. 2.5 [2.6], p = 0.30), higher MAC-Q (26.0 [3.0] vs. 24.0 [3.0], p < 0.001) and HDRS (26.5 [6.0] vs. 24.0 [6.0], p < 0.001), lower emotional stability (48.0 [17.0] vs. 55.0 [1.0], p = 0.015), and a lower score in social activities (7.8 [4.7] vs. 9.6 [5.0], p = 0.030) compared to patients without worry.

As expected, women in menopause were older than women not in menopause (median = 63.1 vs. 49.2 years, p < 0.001, η² = 0.24). After age-adjustment, the only significant difference was the proportion of depressive symptoms, as 25% of non-menopausal women had depressive symptoms (25% vs. 9%, χ² = 5.2, p-_adj_ = 0.022, V = 0.152). There were no other differences between these two groups in any other demographic, clinical, personality or neuropsychological variables.

Patients with CSVD were older than patients without. There were no other differences between these two groups after age-adjustment. There were no differences: between *APOE* ε4 carriers and non-carriers (except for the difference between men and women, as showed above); between women that used HRT and women who did not (except for the difference in sleep disorder, as showed above); between patients with family history of dementia and patients without.

#### 3.1.8 Features associated with NPS scores

As shown above, some NPS tasks were associated with more than one variable. To investigate which variables independently influenced NPS performances, we performed multivariate linear regression models (Supplementary Table 1). All the models were also adjusted for age at baseline and years of education. We found that the presence of depressive symptoms was the only significant variable in the multivariate model for MMSE. Use of benzodiazepine and depressive symptoms were the two variables associated with constructional praxis, while benzodiazepine use, and anticholinergic use were associated with attention performances. Executive function was associated with depressive symptoms and presence of insomnia. Finally, gender was the only determinant of performances in spatial span and in a language test.

#### 3.1.9 Network analysis

To better understand the relationship between gender, severity of SCD, depressive symptoms, insomnia and use of drugs, we constructed a pairwise Markov random-field (MRF) model considering the following features as nodes: age at onset, gender, symptom duration, years of education (education), TIB, HDRS, MAC-Q, insomnia, use of benzodiazepines, and use of anticholinergics.

Robustness analyses of the edges and the centrality indices (see Methods) showed the following Cs-coefficients: 0.516 for edges, 0.516 for strength, 0.205 for closeness, and 0.05 for betweenness. Only edges and strength can then be considered stable [42].

We investigated the connection between gender, TIB score (as a cognitive reserve index), age at onset, MAC-Q score (as an estimation of SCD severity) and HDRS score. The estimated network (Figure 1.A) showed an inverse connection between female gender and the score at TIB, with a weight significantly stronger than all the other edges, except for the connection between MAC-Q and the use of anticholinergics (Supplementary Figure 1). As expected, TIB and years of education were strongly directly connected, with a weight significantly higher than all the other edges, except for the connections between BZD and anticholinergic use, and between BZD use and MAC-Q. Age at onset was inversely connected with symptom duration, education, and MAC-Q, with no differences among the weights of the three edges. MAC-Q was directly connected with sleep disorder, use of BZD, symptom duration, and female gender, with no differences among these edges, while it was inversely and equally connected with the use of anticholinergics, age at onset, and education. HDRS was directly and equally (with no significant differences) connected with education, female gender, the use of anticholinergics, and sleep disorder.

**Figure 1.**
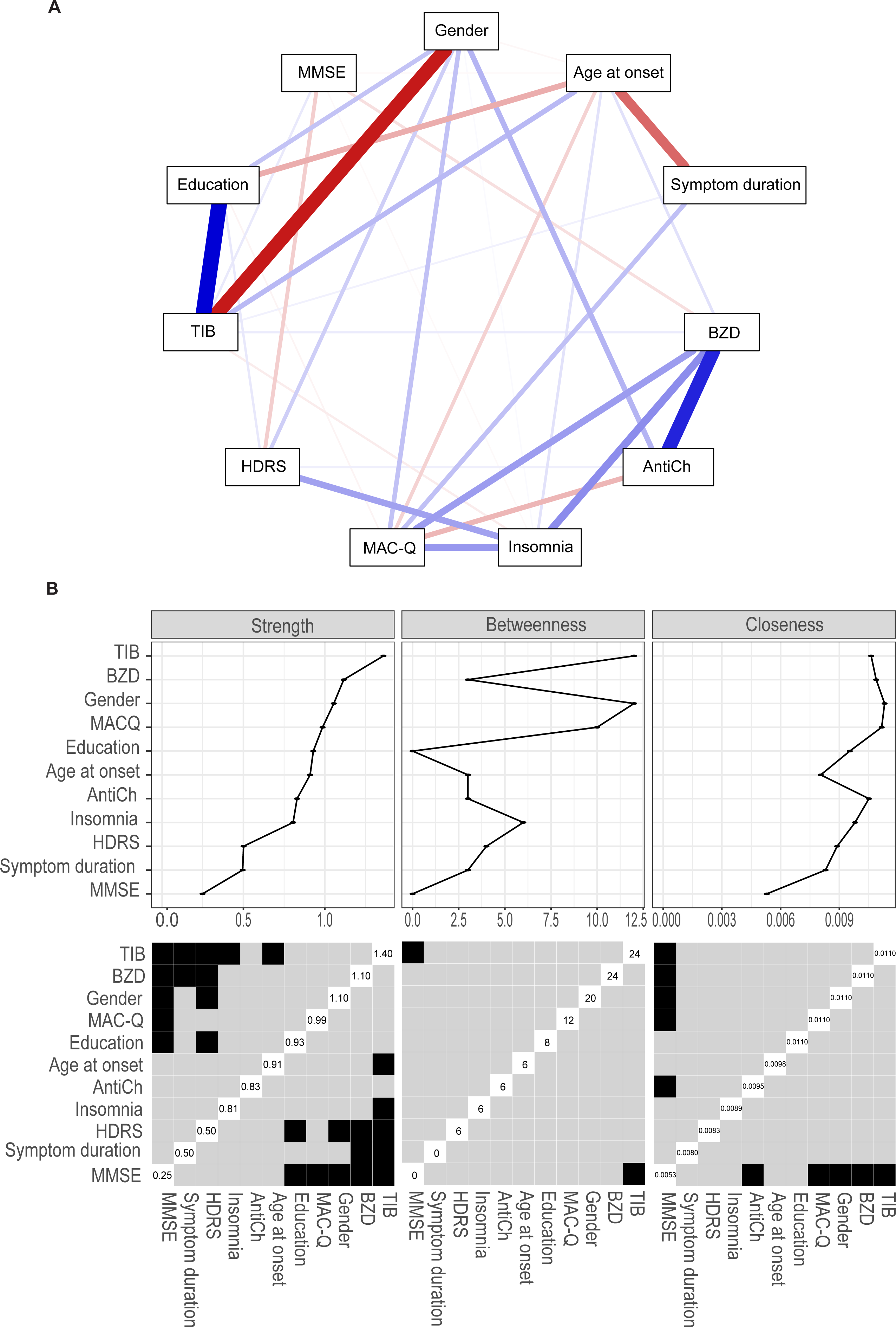
Estimated network (A) and centrality indexes (strength, betweenness and closeness) and differences in centrality indices among features (B). **A:** Nodes (features) are represented in rectangles and links between nodes (edges) are represented as lines. Each edge represents the interconnections between two features after conditioning on all of the other features. Thickness of edges indicate weight of edges in the network. Blue lines indicate positive inter-connections. Red lines indicate negative inter-connections. The plot was arranged in a circular layout. **B:** Strength indicates which node has the strongest overall connection. Betweenness measures the number of times a node lies on the shortest path between two other nodes. Closeness summarizes the average distance of a node to all other nodes in the network. Black boxes indicate significant differences, gray boxes indicate non-significant differences. The values in the boxes along the diagonals indicate the strength, closeness, and betweenness values for each variable.

The estimated network did not show connection between HDRS and MAC-Q or between HDRS and age at onset. MMSE had poor connection, directly with TIB and inversely only with HDRS, BZD use and sleep disorder, with no differences between edge weights. Significant differences among edges are summarized in Supplementary Figure 1.

Regarding centrality indexes, TIB, gender and use of benzodiazepine had a higher strength compared to HDRS and symptom duration (Figure 1.B). TIB also had a higher strength than age at onset. TIB and gender had a higher closeness than HDRS, and MAC-Q. There were no significant differences among the considered features regarding betweenness.

### 3.2 Longitudinal analysis

226 patients (159 [70.3%] women and 67 [29.7%] men) had at least one follow-up visit after one year (Figure 2), with a median follow-up time of 9.7 years (IQR = 10.3). Among these, 136 patients (94 [69.1%] women and 42 [30.9%] men) remained cognitively unimpaired. This group included: 13 patients (nine women and four men) diagnosed with depression (one of them reverted to absence of cognitive complaint after therapy with SSRI); three women diagnosed as affected by OSAS; one woman with vitamin B12 deficit whose SCD reverted to absence of cognitive complaint after B12 correction. None of the 12 patients diagnosed with depression were taking SSRI/SNRI or BZD before the diagnosis and only two patients had subthreshold depressive symptoms at baseline. For 119 patients with stable SCD, we did not identify any potential underlying cause. We defined them as SCD of unknown cause.

**Figure 2.**
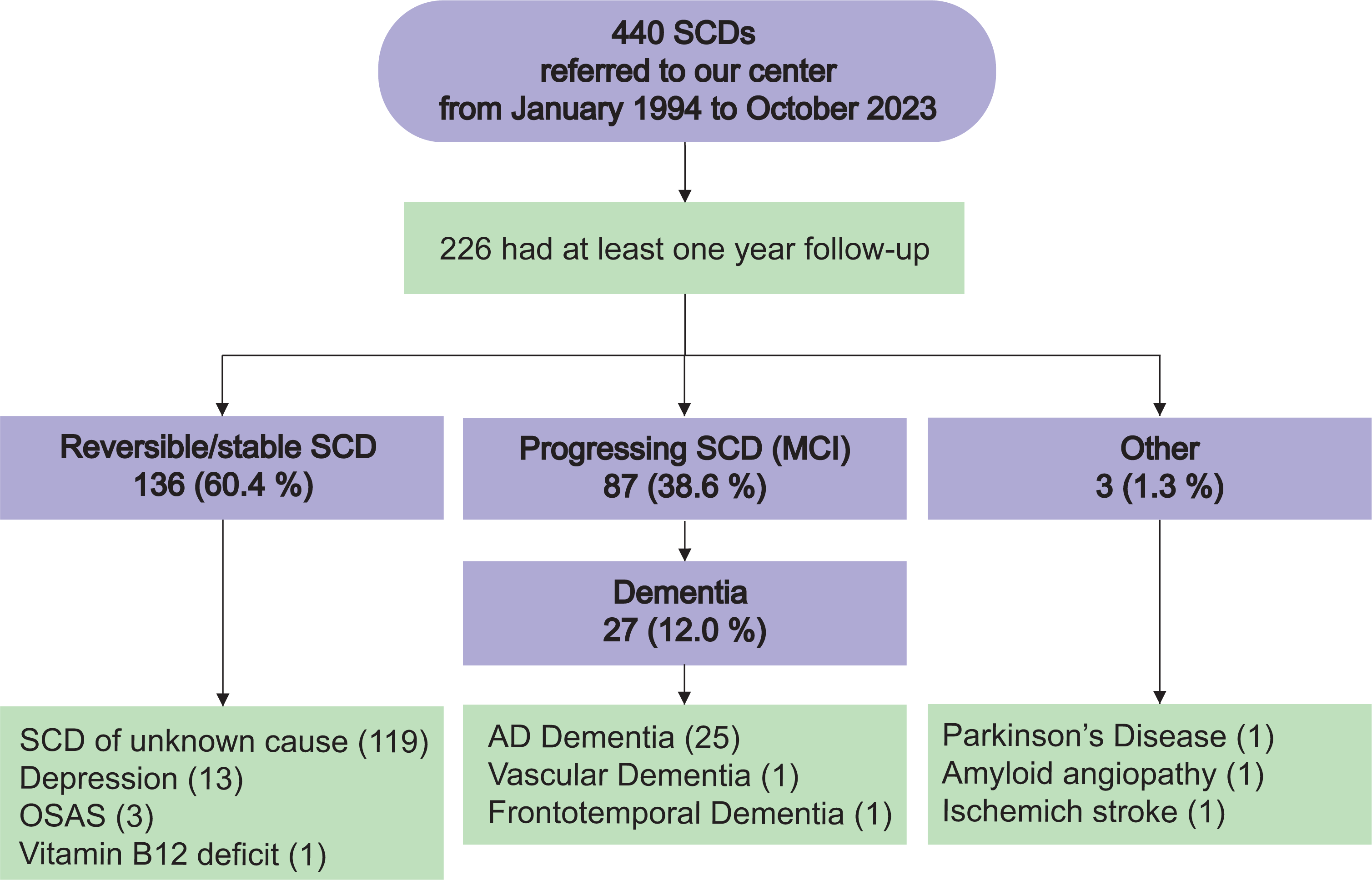
Different trajectories of SCD during the follow-up.

Eighty-seven patients (63 [72.4 %] women and 24 [27.6 %] men) showed progression to objective cognitive decline (MCI) and were classified as progressing SCD (p-SCD). Among these, 27 (18 women and nine men) patients furtherly progressed to dementia, including 25 patients with AD, one with vascular dementia (VaD, with negative AD biomarkers) and one patient with frontotemporal dementia (FTD, started as non-fluent primary progressive aphasia). Finally, three patients developed a neurological disease without dementia (Parkinson disease, ischemic stroke and hemorrhagic stroke due to amyloid angiopathy).

Three patients who were younger than 40 at onset progressed to MCI. One of them progressed to AD dementia (a woman with estimate onset of SCD at her 23, she had the first neurological assessment at 49 and developed dementia when she was 57 years old); another one (a woman with onset at 33 and first evaluation at 49) had positive Amyloid-PET performed when she was 51. None of the patients who had the first neuropsychological visit before 49 years developed MCI.

#### 3.2.1 Incidence rate of MCI and dementia

The median progression time from SCD to MCI was 7.2 (IQR = 5.2) years (range: 1.0 – 20.0), with a mode at two years. Median progression time from SCD to dementia was 10.8 (IQR = 8.7) years (range: 3.4 – 22.1) with two modes at 6, and 16 years. Based on follow-up time of each patient, we calculated the IR per person-year that was: 4.3% (95% C.I. = 3.5 : 5.3) for progression to MCI and 1.2 % (95% C.I. = 0.8 : 1.7) for conversion to AD (Table 2).

**Table 2.** Descriptive statistics of progression time to MCI and conversion time to AD.

|  | Progression time to<br>MCI | Conversion to<br>dementia |
| --- | --- | --- |
| Mean (SD) | 8.1 (5.2) | 11.0 (4.9) |
| Median (IQR) | 7.2 (5.2) | 10.8 (8.7) |
| Mode | 2 | 6, 16 |
| Min:max | 1.0 : 20.0 | 3.4 : 22.1 |
| IR, % (95% C.I.) | 4.3 (3.5 : 5.3) | 1.2 (0.8 : 1.7) |

Progression and conversion time are expressed as years. IR: incidence rate per person-year

We found no association between time of progression to MCI and conversion to AD with any demographic, clinical, genetic or personality variable.

To explore how the rate of progression to MCI was distributed along the follow-up period, we calculated CIR per each year (Supplementary Figure 2). We identified a first peak of incidence within two years (11%) a second peak at 8 years (16%) and the highest incidence after 17 years.

Based on this distribution, we hypothesized the existence of three population of patients that we defined as: early-progressor (within two years), mean-progressor (within 15 years), late-progressor (after 15 years). We split the group of p-SCD patients according to this classification and tested for differences between these three groups. There were no differences regarding any of the demographic, clinical, genetic, personality and neuropsychological variables, except for scores at Trail Making Test B-A (early-progressors < late-progressors, p = 0.034) and ROCFR (early-progressors < late-progressors, p = 0.004; mean-progressor < late-progressors, p = 0.04).

#### 3.2.2 Comparison between reversible/stable and progressing SCD

For the purposes of this analysis, we considered two groups: i) p-SCD patients, including 87 patients; ii) patients with reversible/stable SCD with at least 10 years of follow-up (r/s-SCD), including 56 patients. We selected this group of patients to reduce the underestimation of progression to objective cognitive impairment in patients with a too short follow-up time. None of the patients included in this analysis was younger than 40 at the baseline assessment (min:max = 42 : 86 years).

We compared p-SCD and r/s-SCD (Table 3): p-SCD were older, had lower score in MMSE and higher emotional stability at baseline, had a higher proportion of *APOE* ε4 and use of BZD, SSRI/SNRI and anticholinergic drugs than r/s-SCD.

**Table 3.** Comparison between r/s-SCD and p-SCD.

| **Demographics** | **r/s-SCD (56)** | **p-SCD (87)** | **p** |
| --- | --- | --- | --- |
| Age at onset | 52.0 (11.0) | 62.0 (10.0) | <u>< 0.001</u> |
| Age at onset > 60 | 12/53 (21.4) | 51/87 (58.6) | <u>< 0.001</u> |
| Age at 1° visit | 58.1 (12.0) | 65.18 (10.0) | <u>< 0.001</u> |
| Symptoms duration | 3.8 (5.0) | 2.8 (2.3) | 0.199 |
| Follow-up time | 15.6 (6.0) | 12.6 (9.4) | <u>0.001</u> |
| Women | 36/53 (64.3) | 63/87 (72.4) | 0.188 |
| Menopause age | 51.0 (6.0) | 50.0 (8.0) | 0.790 |
| Family history of dementia | 30/56 (53.6) | 52/87 (59.8) | 0.595 |
| Confirmation by caregiver | 3/6 | 10/14 | 0.357 |
| Memory complaint | 46/48 (95.8) | 62/64 (96.9) | 0.735 |
| Worry about SCD | 24/30 (80.0) | 29/39 (74.4) | 0.409 |
| **Clinical** |  |  |  |
| Women with use of HRT | 10/36 (27.8) | 14/63 (22.2) | 0.413 |
| Early menopause | 0/33 (0) | 2/62 (3.2) | 0.311 |
| Sleep disorder | 27/57 (47.4) | 48/85 (56.5) | 0.224 |
| HDRS | 3.0 (6.0) | 4 (6.0) | 0.959 |
| Depressive symptoms | 6/49 (12.2) | 4/76 (5.3) | 0.160 |
| MMSE | 30.0 (3.0) | 27.0 (4.0) | <u>< 0.001</u> |
| MAC-Q | 26.0 (2.0) | 26.0 (4.0) | 0.785 |
| APOE ε4 | 7/47 (14.9) | 33/71 (46.5) | <u><0.001</u> |
| **Cognitive reserve** |  |  |  |
| Years of education | 13.0 (7.0) | 11.0 (5.0) | 0.223 |
| TIB | 112.0 (6.3) | 111.9 (9.8) | 0.296 |
| Intellectual activities | 18.5 (5.4) | 17.8 (8.0) | 0.755 |
| Social activities | 8.8 (5.0) | 8.6 (4.6) | 0.399 |
| Physical activities | 5.0 (4.6) | 5.7 (4.3) | 0.482 |
| **Drugs** |  |  |  |
| BZD use | 4/57 (7.0) | 16/85 (18.8) | <u>0.027</u> |
| SSRI/SNRI use | 3/53 (5.7) | 16/85 (18.8) | <u>0.027</u> |
| Anticholinergic use | 8/57 (14.0) | 22/85 (26.2) | <u>0.036</u> |
| **Cardiovascular features** |  |  |  |
| Hypertension | 16/55 (29.1) | 27/84 (32.1) | 0.831 |
| Dyslipidemia | 8/55 (14.5) | 25/84 (29.8) | 0.053 |
| IFG or diabetes | 5/55 (9.1) | 7/81 (8.6) | 0.848 |
| Diabetes | 3/55 (5.5) | 3/81 (3.7) | 0.575 |
| Previous or current smoking habit | 17/56 (30.4) | 29/82 (35.4) | 0.598 |
| Current smoking habit | 5/56 (8.9) | 9/82 (10.9) | 0.756 |
| CSVD | 23/40 (57.5) | 40/61 (65.6) | 0.286 |
| **Personality traits** |  |  |  |
| Energy | 45.0 (12.0) | 44.0 (11.0) | 0.579 |
| Friendship | 50.0 (13.0) | 49.0 (15.0) | 0.265 |
| Conscientiousness | 47.0 (15.0) | 48.0 (10.0) | 0.366 |
| Emotional Stability | 46.5 (16.0) | 54.0 (16.0) | <u>0.008</u> |
| Openness | 45.0 (16.0) | 46.0 (18) | 0.886 |

Values quoted in table are median (IQR) or frequencies (percentages). Groups were compared between cohorts using Mann-Whitney U test or Chi-square tests. Statistically significant cohort group differences ( $p < 0.05$ ) are in underlined characters. HDRS: Hamilton Depression Rating Scale; MAC-Q: Memory Assessment Clinics – Questionnaire; MMSR: MiniMental State Examination; TIB: *Test di Intelligenza Breve*; APOE: apolipoprotein E; SSRI/SNRI: selective serotonin reuptake inhibitor/serotonin and noradrenalin reuptake inhibitor; IFG: impaired fasting glucose; CSVD: cerebral small vessel disease.

p-SCD also had lower scores in neuropsychological task for ecological memory (RMBT, p = 0.021), short-term memory (digit span, p = 0.007), and constructional praxis (CDT, p = 0.047). Among these, only digit span (B = -0.43, p-_adj_ = 0.018) remained significantly different between the two groups after age- and education-adjustment.

We ran a backward logistic regression analysis considering the classification as r/s-SCD or p-SCD as dependent variables and age at 1° visit, age at onset, MMSE, *APOE* ε4, emotional stability, depressive symptoms, use of BZD, us of SSRI/SNRI and use anticholinergics covariates. The final model was statistically significant (χ² = 31.9, p-_adj_ < 0.001) and included as significant covariates: age at onset (B = 0.07, p-_adj_ = 0.031), MMSE (B = -0.30, p-_adj_ = 0.041), *APOE* ε4 (B = 1.83, p-_adj_ = 0.010) and emotional stability (B = 0.062, p-_adj_ = 0.032). Use of SSRI/SNRI was include in the model but did not reach the statistical significance (B = 2.00, p-_adj_ = 0.071).

#### 3.2.3 Machine learning model

The XGBoost model with optimized hyperparameters achieved a balanced accuracy of 74.37% [66.97 : 80.90]. Other performance metrics are reported in Supplementary Table 2 and Supplementary Figure 3. Average absolute SHAP values for each feature are reported in Figure 3A, while single subject SHAP values for the top 20 features are depicted in Figure 3B. The significantly different comparisons between feature importances after Bonferroni correction are shown in Figure 3C and their raw p-values in Supplementary Figure 4.

**Figure 3.**
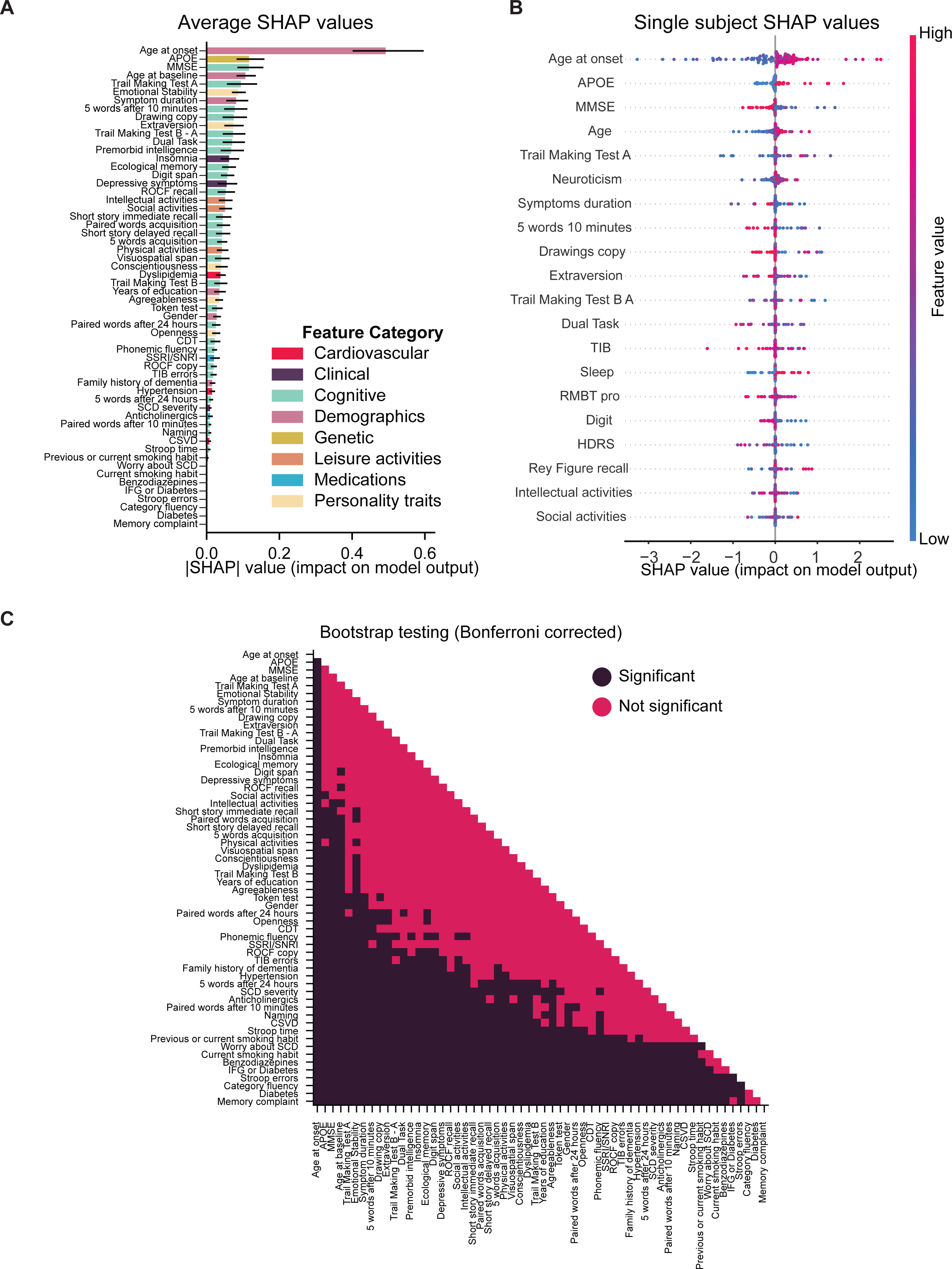
Feature importance based on SHAP values. **A**: Mean absolute value of the mean SHAP value for each feature. Error bar indicates 95% confidence interval obtained by bootstrapping (n = 10e3). Color labels indicate different domains of the input variable. **B**: SHAP values, divided by feature, for each subject for the top 20 features. Each dot indicates a single subject, and the colorbar depicts the (min-max normalized) value of the feature. **C**: Significance of the bootstrap test for pairwise comparison between feature importances. Each square represents the binary significance of the comparison between two feature importance weights. Darker squares indicate significant differences between the two features, lighter squares indicate that the difference is not significant (alpha = 0.05, Bonferroni corrected).

#### 3.2.4 Amyloid and neurodegeneration biomarker analysis

Among patients who underwent CSF or Amyloid PET, 33.3% were A+. The majority of them (16.2%) had a A+/T+/N+ profile while only two patients had a A+/T+/N- profile. Three patients (8.1%) had a A-/T-/N+ profile. Only one patient had a A-/T+/N+ profile (Figure 4). There were no differences regarding any considered (demographic, clinical or cognitive) feature between A- and A+ patients. Concentrations of CSF biomarker and proportions of positivity for each biomarker are shown in Supplementary Table 3.

**Figure 4.**
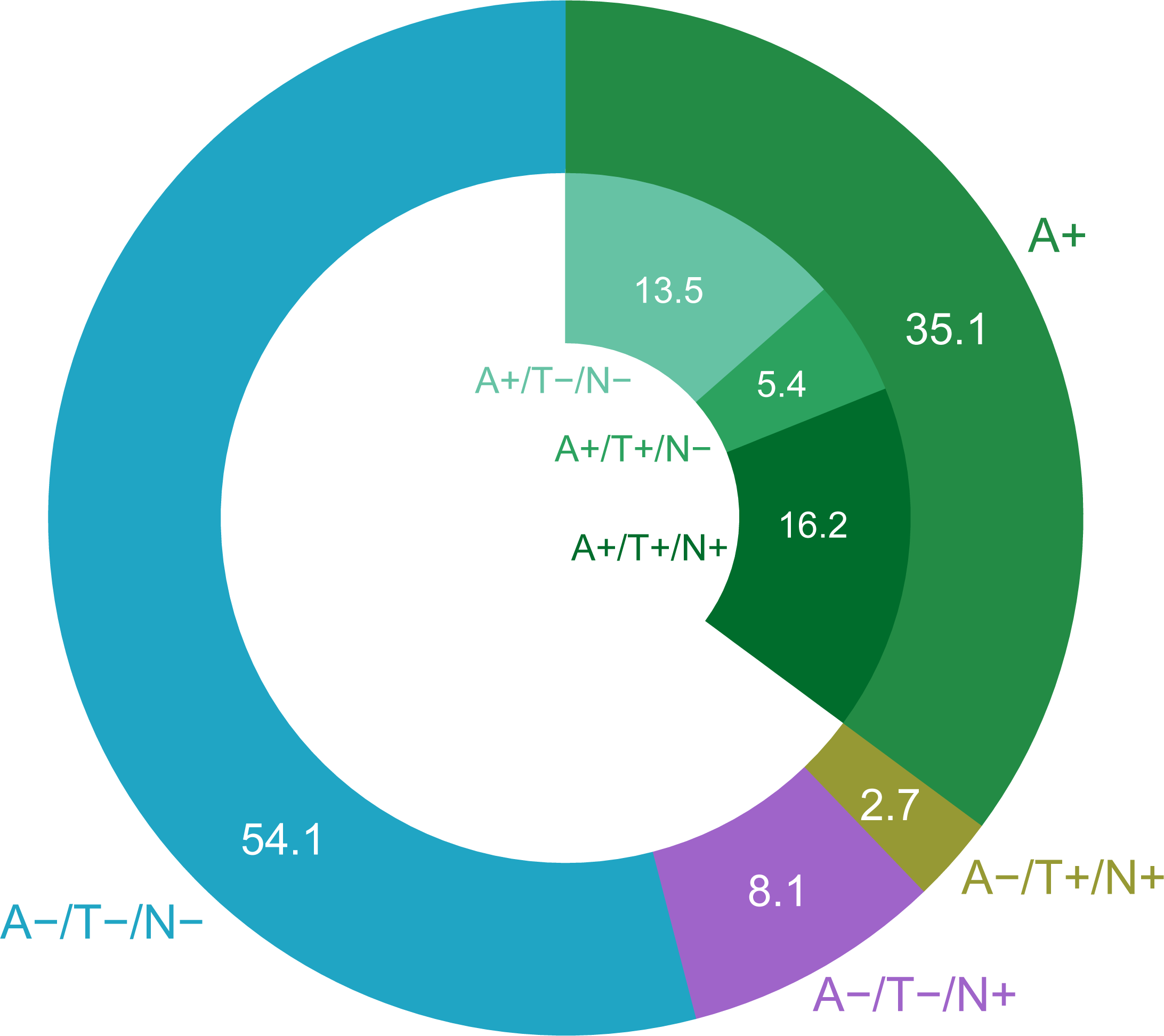
Donut plot showing proportions of biomarker profile in SCD patients. Values plotted in the donut are percentages. The proportion of A+ patients referred to the group of 37 patients who underwent CSF.

#### 3.2.5 Longitudinal analysis of the subgroup with AD biomarker assessment

Patients who underwent AD biomarker assessment (CSF or Amyloid-PET) had a median follow-up time of 2 years (IQR 0.7, min:max = 0.2 : 9.6). During this period, ten patients progressed to MCI (p-SCD) in a median time of 2.3 (IQR = 1.5, min:max = 1.4 : 5.7). Patients who did not progress had a median follow-up time of 2.0 (IQR = 1.3, min:max = 0.2 : 9.6). No patient progressed to dementia. Among p-SCD patients, eight (80.0 %) were A+ at baseline. Six out of 14 (42.6%) patients who were A+ did not progress during a median follow-up time of 3.2 years (IQR = 1.9, min:max = 1.4 : 5.7). Twenty-seven out of 29 (93.1 %) patients who were A-, did not progress to MCI. To be noted, among the two p-SCD patient who were A-, one had a CSF Aβ_42_/Aβ_40_ of 0.066 (cut-off < 0.062) and was T+ and N+, but did not underwent amyloid-PET. The other one only underwent amyloid-PET and did not undergo CSF biomarker assessment.

## 4 Discussion

### 4.1 Characteristics of SCD population

Demographic characteristics and *APOE* ε4 proportions in our sample align with previous memory studies [49,50]. Women constituted more than twice the number of men, consistent with prior research [51], and exhibited younger age, lower education levels, more severe SCD, and higher rates of depressive symptoms and sleep disorders. Conversely, men had a higher proportion of *APOE* ε4, suggesting that SCD in women may be linked to reversible causes (mood or sleep disturbances), while in men it may be more associated with underlying pathological processes.

The age of onset in SCD spans a broad range (34–86 years). Patients over 60 showed a higher rate of cognitive decline progression, more *APOE* ε4 carriers, and a higher prevalence of factors that could influence cognitive decline, such as lower cognitive reserve [52,53] and a greater incidence of hypertension and CSVD [54]. Thus, older patients clearly require closer monitoring for potential cognitive decline progression, while younger patients are more likely to experience treatable conditions that may adversely affect their quality of life [21]. Among clinical features, insomnia and mild CSVD affected up to 60% of the sample, while other conditions, such as early menopause, depressive symptoms, diabetes, or smoking, were present in less than 10% of patients.

### 4.2 Factors influencing cognitive performance and SCD presentation

Cognitive performance, specifically in constructional praxis, attention, executive function, and auditory comprehension, was associated with gender, depressive symptoms, insomnia, and the use of benzodiazepines or anticholinergic drugs. These neuropsychological functions, however, are not associated with future progression to MCI or dementia [55], indicating that slight variations in scores may reflect modifiable conditions present at baseline rather than a risk of cognitive decline. To explore the relationships between SCD severity, gender, depressive symptoms, and cognitive reserve, we constructed a Markov random field network. Two primary poles emerged in the network: one connecting gender, cognitive reserve, age at onset, and symptom duration, and another linking SCD severity with medication use and insomnia. Interestingly, depressive symptoms were not connected with SCD severity or age at onset. Indeed, “depressive symptoms” was the least central feature in the network. However, it was a significant predictor of MMSE, executive function, and constructional praxis scores. This suggests that while depressive symptoms may not drive the onset or severity of SCD, they influence the SCD cognitive profile. This supports prior findings of a synergistic relationship between SCD and depressive symptoms, rather than a direct cause-effect link [51].

Benzodiazepine use had a notable impact on SCD severity, consistent with earlier studies [56]. As expected, benzodiazepine use is also strongly associated with sleep disorders. We are unable to establish a clear causal relationship (whether insomnia leads to increased benzodiazepine use and, consequently, greater SCD severity, or if the severity of SCD contributes to both insomnia and increased benzodiazepine use). However, it is plausible that insomnia, benzodiazepine use, and SCD severity are interconnected, with each factor mutually influencing and exacerbating the others.

### 4.3 SCD progression trajectories

Progression from SCD to objective cognitive decline is a slow process, with median times to MCI and dementia of 7 and 11 years, respectively. The incidence rates per person-year were 4.3% for MCI and 1.2% for dementia, aligning with previous findings [57,58]. AD was the most common diagnosis among those who progressed to dementia, as previously reported [59], with only one patient diagnosed with vascular dementia and another with non-fluent/agrammatic variant of primary progressive aphasia. In our sample, the most common non-degenerative causes of SCD was depression, followed by obstructive sleep apnea syndrome (OSAS), and vitamin B12 deficiency, all of which are recognized contributors to cognitive impairment also in literature [60– 62]. However, in the majority of cases, no underlying cause was identified, leading us to classify these patients as having SCD of unknown cause. Three cases of progression to MCI, and even early-onset Alzheimer’s disease, were observed in patients with early age of onset (23 years). Although it is challenging to definitively prove that SCD reported by individuals in their twenties could represent the first signs of an underlying degenerative process, the symptom duration and time to disease progression in these cases are consistent with the timeframe required for Aβ deposition to advance to cognitive impairment. Hence, SCD in young individuals deserves attention, especially as we enter the era of DMTs, to identify patients who may benefit from early intervention.

### 4.4 Predictors of progression to MCI and dementia

Our machine learning model predicted progression from SCD to MCI and dementia with good accuracy and provided a hierarchical ranking of the contributing variables. Interestingly, only a small subset of features is completely discarded (SHAP-value 0), while the rest are all used, albeit in very different proportions. The most prominent result was the importance of age at onset, surpassing all other variables, followed by *APOE* ε4, confirming previous researches [25,55,63]. Neuropsychological tests (evaluating memory and executive functions), personality traits, and cognitive reserve proxies emerged as some of the most important predictors, confirming previous findings [55,64], and significantly outweighing other variables. Insomnia emerged as the most significant clinical predictor, corroborating earlier findings [65].

Despite the influence of gender on SCD, its role in progression to dementia was minimal. Similarly, depressive symptoms and the use of benzodiazepines or anticholinergics were less important. This suggests that while these factors contribute to SCD’s presentation, they may not be linked to cognitive decline progression.

Previous research showed that greater white matter signal abnormalities and elevated cardiovascular risk are associated with increased likelihood of amyloid burden and progression to dementia [54,66]. In our cohort, metabolic and cardiovascular comorbidities did not significantly impact SCD, though it is important to note that the prevalence of these conditions was lower compared to other studies [49], likely due to the inclusion of younger participants.

### 4.5 Prevalence of AD in SCD population

More than one-third of patients were A+, a proportion that exceeds the reported prevalence of abnormal Aβ in populations without SCD [67,68]. Furthermore, we observed distinct trajectories in SCD patients based on their biomarker status: 80% of those who progressed to MCI were A+, with the two remaining patients potentially representing false negatives. Only two out of 29 patients with negative AD biomarkers progressed to MCI. Although our subsample size is small, these data may indicate that progression from SCD to MCI is, in most cases, suggestive of future progression to AD.

### 4.6 Approach to managing SCD patients

We propose an operational protocol (Figure 5), based on the findings discussed in previous sections and currently implemented in our memory clinic (Supplementary Figure 5). We distinguish between clinical and research settings and define specific time points: baseline, short-term follow-up, and long-term follow-up. For the baseline assessment, we provide a comprehensive list of features to evaluate and tests to perform. The short-term follow-up period should focus on the timeframe during which a patient is most likely to progress, determined by their risk status. Based on our findings of multiple peaks in MCI incidence, we recommend short-term follow-up intervals of two to five years. Long-term follow-up is the phase where, depending on the patient’s trajectory, decisions can be made to either discharge the patient or conduct further evaluations. We identify three potential trajectories:

**Figure 5.**
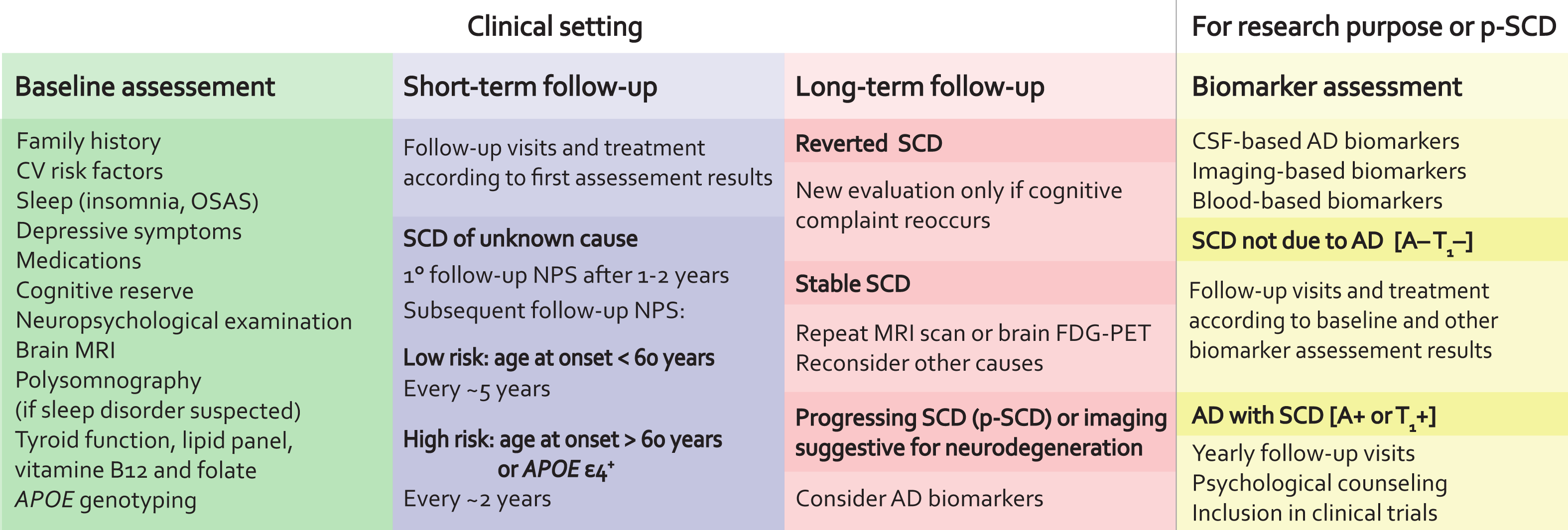
Proposal for a management protocol of SCD. The proposal is divided into two main components: one for clinical practice and another for research purposes. For the clinical setting, we have identified three key time points: baseline, short-term follow-up, and long-term follow-up. The baseline assessment includes all evaluations and tests that should be conducted at the initial examination of patients. The short-term follow-up management depends on the baseline assessment results: if a potential cause of SCD is identified, the patient should receive appropriate treatment and be followed up accordingly. If no cause is found, we propose classifying the patient as having “SCD of unknown cause” and performing neuropsychological assessments (NPS) after 1-2 years. Subsequent NPS evaluations should be guided by the estimated risk of cognitive decline progression. This risk assessment is based on our machine learning results and prior literature, with the primary determinant being age at onset: low risk if the onset age is < 60 years (follow-up NPS after ∼5 years), and high risk if the onset age is > 60 years or if *APOE* ε4 is present (follow-up NPS after ∼2 years). After this period, it is possible to determine the SCD trajectory: reverted SCD (if symptoms are no longer present), stable SCD (if the patient still reports cognitive decline with no progression in NPS tests), or progressing SCD (if NPS shows worsening cognitive performance or brain imaging indicates neurodegeneration). In the latter case, we recommend considering AD biomarker assessment. In a research context, if an SCD patient undergoes AD biomarker assessment, we suggest applying the diagnostic criteria recently proposed by the Alzheimer’s Association Workgroup.

1. Reverted SCD: the patient no longer reports symptoms of SCD and can be discharged, with recommendation for re-evaluation if symptoms reoccur.
2. Stable SCD: the patient continues to report cognitive decline, but without evidence of worsening in neuropsychological evaluations. Additional tests may be considered: in particular we suggest brain ^18^F-FDG-PET or repeated brain MRI as indicators of neurodegenerations [37].
3. Progressive SCD: the patient shows progression to MCI or exhibits neuroimaging evidence suggestive of neurodegeneration, warranting AD biomarker testing.

In all other cases, we concur with the AA Workgroup, which recommends that AD biomarkers be assessed solely for research purposes [37]. For patients undergoing AD biomarker analysis, we recommend applying the revised criteria for AD [37], classifying SCD patients with positive biomarkers as “AD with SCD” and recommending annual follow-up, psychological support, and potential enrollment in AD therapy trials. Patients with negative biomarkers should be classified as “SCD not due to AD” and managed based on the results of the baseline assessment and any additional biomarkers for other diseases, if available.

### 4.7 Limitations and strengths

Key limitations of our study include a high proportion of missing data, limited availability of AD biomarkers, and follow-up data for only half of the sample. Some clinical variables were obtained anamnestically, and CSVD was evaluated via visual inspection. Moreover, an independent validation sample and a control group of cognitively healthy individuals are lacking. Despite these limitations, the study’s strengths include a comprehensive set of variables covering multiple aspects of SCD and extensive follow-up. Moreover, this is the first study that provides a clinical-care guideline for the management of patients with SCD.

### 4.8 Conclusions

We detailed characteristics of SCD, and their impact on cognitive decline progression. Based on these findings, we proposed a protocol for managing SCD that not only focuses on detecting neurodegenerative conditions but also addresses non-degenerative causes. Given the widespread nature of SCD, we emphasize the importance of tailored monitoring based on individual risk factors. This work has a clinical and care-related impact on the management of SCD, laying the groundwork for developing guidelines for its management.

## Abbreviations

SCD: Subjective Cognitive Decline
NPS: Neuropsychological tests
FDG-PET: Fluorodeoxyglucose Positron Emission Tomography
FBB: 18F Florbetaben
FMM: 18F-Flutemetamol
NIA-AA: National Institute on Aging-Alzheimer’s Association
IQR: Interquartile Range
BZD: Benzodiazepine
HRT: Hormone Replacement Therapy
IFG: Impaired Fasting Glucose
CSVD: Cerebral Small Vessel Disease
MAC-Q: Memory Assessment Clinics-Questionnaire
HDRS: Hamilton Depression Rating Scale
TIB: Test di Intelligenza Breve
OSAS: Obstructive Sleep Apnea Syndrome
VaD: Vascular Dementia
IR: Incidence Rate

## Data Availability

All data produced in the present study are available upon reasonable request to the authors

## Consent statement

All participants in this study provided informed consent, agreeing to participate and to share the results deriving from their data.

## References

[1] Turvey CL, Schultz S, Arndt S, Wallace RB, Herzog R. Memory complaint in a community sample aged 70 and older. J Am Geriatr Soc 2000;48:1435–41.

[2] Paradise MB, Glozier NS, Naismith SL, Davenport TA, Hickie IB. Subjective memory complaints, vascular risk factors and psychological distress in the middle-aged: a cross-sectional study. BMC Psychiatry 2011;11:108. 10.1186/1471-244X-11-108.

[3] Jonker C, Geerlings MI, Schmand B. Are memory complaints predictive for dementia? A review of clinical and population-based studies. Int J Geriatr Psychiatry 2000;15:983–91. 10.1002/1099-1166(200011)15:11<983::aid-gps238>3.0.co;2-5.

[4] Abdulrab K, Heun R. Subjective Memory Impairment. A review of its definitions indicates the need for a comprehensive set of standardised and validated criteria. Eur Psychiatry 2008;23:321–30. 10.1016/j.eurpsy.2008.02.004.

[5] O’Connor DW, Pollitt PA, Roth M, Brook PB, Reiss BB. Memory complaints and impairment in normal, depressed, and demented elderly persons identified in a community survey. Arch Gen Psychiatry 1990;47:224–7.

[6] Bolla KI, Lindgren KN, Bonaccorsy C, Bleecker ML. Memory complaints in older adults. Fact or fiction? Arch Neurol 1991;48:61–4. 10.1001/archneur.1991.00530130069022.

[7] Grut M, Jorm AF, Fratiglioni L, Forsell Y, Viitanen M, Winblad B. Memory complaints of elderly people in a population survey: variation according to dementia stage and depression. J Am Geriatr Soc 1993;41:1295–300. 10.1111/j.1532-5415.1993.tb06478.x.

[8] Jessen F, Amariglio RE, van Boxtel M, Breteler M, Ceccaldi M, Chételat G, et al. A conceptual framework for research on subjective cognitive decline in preclinical Alzheimer’s disease. Alzheimer’s & Demential: The Journal of the Alzheimer’s Association 2014;10:844–52. 10.1016/j.jalz.2014.01.001.

[9] Parfenov VA, Zakharov VV, Kabaeva AR, Vakhnina NV. Subjective cognitive decline as a predictor of future cognitive decline: a systematic review. Dement Neuropsychol 2020;14:248–57. 10.1590/1980-57642020dn14-030007.

[10] Jack CR, Bennett DA, Blennow K, Carrillo MC, Dunn B, Haeberlein SB, et al. NIA-AA Research Framework: Toward a biological definition of Alzheimer’s disease. Alzheimers Dement 2018;14:535–62. 10.1016/j.jalz.2018.02.018.

[11] Mitchell AJ, Beaumont H, Ferguson D, Yadegarfar M, Stubbs B. Risk of dementia and mild cognitive impairment in older people with subjective memory complaints: meta-analysis. Acta Psychiatr Scand 2014;130:439–51. 10.1111/acps.12336.

[12] Margolis SA, Kelly DA, Daiello LA, Davis J, Tremont G, Pillemer S, et al. Anticholinergic/Sedative Drug Burden and Subjective Cognitive Decline in Older Adults at Risk of Alzheimer’s Disease. The Journals of Gerontology: Series A 2020. 10.1093/gerona/glaa222.

[13] Vellas B, Aisen PS, Sampaio C, Carrillo M, Scheltens P, Scherrer B, et al. Prevention trials in Alzheimer’s disease: an EU-US task force report. Prog Neurobiol 2011;95:594–600. 10.1016/j.pneurobio.2011.08.014.

[14] Cummings J, Zhou Y, Lee G, Zhong K, Fonseca J, Cheng F. Alzheimer’s disease drug development pipeline: 2023. Alzheimer’s & Dementia: Translational Research & Clinical Interventions 2023;9:e12385. 10.1002/trc2.12385.

[15] Jack CR, Bennett DA, Blennow K, Carrillo MC, Dunn B, Haeberlein SB, et al. NIA-AA Research Framework: Toward a biological definition of Alzheimer’s disease. Alzheimers Dement 2018;14:535–62. 10.1016/j.jalz.2018.02.018.

[16] Bhome R, Berry AJ, Huntley JD, Howard RJ. Interventions for subjective cognitive decline: systematic review and meta-analysis. BMJ Open 2018;8:e021610. 10.1136/bmjopen-2018-021610.

[17] Roehr S, Luck T, Pabst A, Bickel H, König H-H, Lühmann D, et al. Subjective cognitive decline is longitudinally associated with lower health-related quality of life. Int Psychogeriatr 2017;29:1939–50. 10.1017/S1041610217001399.

[18] Gifford KA, Bell SP, Liu D, Neal JE, Turchan M, Shah AS, et al. Frailty Is Related to Subjective Cognitive Decline in Older Women without Dementia. Journal of the American Geriatrics Society 2019;67:1803–11. 10.1111/jgs.15972.

[19] Taylor CA. Subjective Cognitive Decline Among Adults Aged ≥45 Years — United States, 2015– 2016. MMWR Morb Mortal Wkly Rep 2018;67. 10.15585/mmwr.mm6727a1.

[20] Bubbico G, Di Iorio A, Lauriola M, Sepede G, Salice S, Spina E, et al. Subjective Cognitive Decline and Nighttime Sleep Alterations, a Longitudinal Analysis. Front Aging Neurosci 2019;11. 10.3389/fnagi.2019.00142.

[21] Bouldin ED, Taylor CA, Knapp KA, Miyawaki CE, Mercado NR, Wooten KG, et al. Unmet needs for assistance related to subjective cognitive decline among community-dwelling middle-aged and older adults in the US: prevalence and impact on health-related quality of life. Int Psychogeriatr 2020:1–14. 10.1017/S1041610220001635.

[22] Lawton MP, Brody EM. Assessment of Older People: Self-Maintaining and Instrumental Activities of Daily Living. The Gerontologist 1969;9:179–86. 10.1093/geront/9.3_Part_1.179.

[23] McKhann GM, Knopman DS, Chertkow H, Hyman BT, Jack CR, Kawas CH, et al. The diagnosis of dementia due to Alzheimer’s disease: recommendations from the National Institute on Aging-Alzheimer’s Association workgroups on diagnostic guidelines for Alzheimer’s disease. Alzheimers Dement J Alzheimers Assoc 2011;7:263–9. 10.1016/j.jalz.2011.03.005.

[24] Albert MS, DeKosky ST, Dickson D, Dubois B, Feldman HH, Fox NC, et al. The diagnosis of mild cognitive impairment due to Alzheimer’s disease: recommendations from the National Institute on Aging-Alzheimer’s Association workgroups on diagnostic guidelines for Alzheimer’s disease. Alzheimers Dement 2011;7:270–9. 10.1016/j.jalz.2011.03.008.

[25] Mazzeo S, Padiglioni S, Bagnoli S, Carraro M, Piaceri I, Bracco L, et al. Assessing the effectiveness of subjective cognitive decline plus criteria in predicting the progression to Alzheimer’s disease: an 11-year follow-up study. Eur J Neurol 2020;27:894–9. 10.1111/ene.14167.

[26] Slot RER, Verfaillie SCJ, Overbeek JM, Timmers T, Wesselman LMP, Teunissen CE, et al. Subjective Cognitive Impairment Cohort (SCIENCe): study design and first results. Alzheimer’s Research & Therapy 2018;10. 10.1186/s13195-018-0390-y.

[27] Campbell NL, Boustani MA, Lane KA, Gao S, Hendrie H, Khan BA, et al. Use of anticholinergics and the risk of cognitive impairment in an African American population. Neurology 2010;75:152–9. 10.1212/WNL.0b013e3181e7f2ab.

[28] Fazekas F, Barkhof F, Wahlund LO, Pantoni L, Erkinjuntti T, Scheltens P, et al. CT and MRI rating of white matter lesions. Cerebrovasc Dis 2002;13 Suppl 2:31–6. 10.1159/000049147.

[29] Goldberg LR. The development of markers for the Big-Five factor structure. Psychol Assess 1992;4:26–42. 10.1037/1040-3590.4.1.26.

[30] Colombo L, Brivio C, Benaglio I, Siri S, Capp SF. Alzheimer patients’ ability to read words with irregular stress. Cortex 2000;36:703–14. 10.1016/s0010-9452(08)70547-1.

[31] Bright P, Hale E, Gooch VJ, Myhill T, van der Linde I. The National Adult Reading Test: restandardisation against the Wechsler Adult Intelligence Scale-Fourth edition. Neuropsychol Rehabil 2018;28:1019–27. 10.1080/09602011.2016.1231121.

[32] Hamilton M. A rating scale for depression. J Neurol Neurosurg Psychiatr 1960;23:56–62.

[33] Crook TH, Feher EP, Larrabee GJ. Assessment of memory complaint in age-associated memory impairment: the MAC-Q. Int Psychogeriatr 1992;4:165–76. 10.1017/s1041610292000991.

[34] Yarnold PR, Stille FC, Martin GJ. Cross-sectional psychometric assessment of the Functional Status Questionnaire: use with geriatric versus nongeriatric ambulatory medical patients. Int J Psychiatry Med 1995;25:305–17. 10.2190/GP4F-WQK9-WRHY-7JM9.

[35] Sorbi S, Nacmias B, Forleo P, Latorraca S, Gobbini I, Bracco L, et al. ApoE allele frequencies in Italian sporadic and familial Alzheimer’s disease. Neurosci Lett 1994;177:100–2. 10.1016/0304-3940(94)90054-x.

[36] Minoshima S, Drzezga AE, Barthel H, Bohnen N, Djekidel M, Lewis DH, et al. SNMMI Procedure Standard/EANM Practice Guideline for Amyloid PET Imaging of the Brain 1.0. J Nucl Med 2016;57:1316–22. 10.2967/jnumed.116.174615.

[37] Jack Jr. CR, Andrews JS, Beach TG, Buracchio T, Dunn B, Graf A, et al. Revised criteria for diagnosis and staging of Alzheimer’s disease: Alzheimer’s Association Workgroup. Alzheimer’s & Dementia 2024;20:5143–69. 10.1002/alz.13859.

[38] Jack CR, Bennett DA, Blennow K, Carrillo MC, Dunn B, Haeberlein SB, et al. NIA-AA Research Framework: Toward a biological definition of Alzheimer’s disease. Alzheimers Dement 2018;14:535–62. 10.1016/j.jalz.2018.02.018.

[39] Epskamp S, Fried EI. A tutorial on regularized partial correlation networks. Psychol Methods 2018;23:617–34. 10.1037/met0000167.

[40] Epskamp S, Cramer AOJ, Waldorp LJ, Schmittmann VD, Borsboom D. qgraph: Network Visualizations of Relationships in Psychometric Data. Journal of Statistical Software 2012;48:1–18. 10.18637/jss.v048.i04.

[41] Papachristou N, Barnaghi P, Cooper BA, Hu X, Maguire R, Apostolidis K, et al. Congruence Between Latent Class and K-Modes Analyses in the Identification of Oncology Patients With Distinct Symptom Experiences. J Pain Symptom Manage 2018;55:318–333.e4. 10.1016/j.jpainsymman.2017.08.020.

[42] Epskamp S, Borsboom D, Fried EI. Estimating psychological networks and their accuracy: A tutorial paper. Behav Res Methods 2018;50:195–212. 10.3758/s13428-017-0862-1.

[43] Chen T, Guestrin C. XGBoost: A Scalable Tree Boosting System. Proceedings of the 22nd ACM SIGKDD International Conference on Knowledge Discovery and Data Mining, 2016, p. 785–94. 10.1145/2939672.2939785.

[44] Chawla NV, Bowyer KW, Hall LO, Kegelmeyer WP. SMOTE: Synthetic minority over-sampling technique. Journal of Artificial Intelligence Research 2002;16:321–57. 10.1613/jair.953.

[45] Bentéjac C, Csörgő A, Martínez-Muñoz G. A Comparative Analysis of XGBoost. Artif Intell Rev 2021;54:1937–67. 10.1007/s10462-020-09896-5.

[46] Cawley GC, Talbot NLC. On Over-fitting in Model Selection and Subsequent Selection Bias in Performance Evaluation. J Mach Learn Res 2010;11:2079–107.

[47] M H, M.N S. A Review on Evaluation Metrics for Data Classification Evaluations. IJDKP 2015;5:01–11. 10.5121/ijdkp.2015.5201.

[48] Lundberg S, Lee S-I. A Unified Approach to Interpreting Model Predictions 2017. 10.48550/arXiv.1705.07874.

[49] Yang K, Chen G, Sheng C, Xie Y, Li Y, Hu X, et al. Cognitive Reserve, Brain Reserve, APOEl4, and Cognition in Individuals with Subjective Cognitive Decline in the SILCODE Study. J Alzheimers Dis 2020;76:249–60. 10.3233/JAD-200082.

[50] Huang Y, Xue J, Li C, Chen X, Fu H, Fei T, et al. Depression and APOEε4 Status in Individuals with Subjective Cognitive Decline: A Meta-Analysis. Psychiatry Investig 2020;17:858–64. 10.30773/pi.2019.0324.

[51] Hao L, Sun Y, Li Y, Wang J, Wang Z, Zhang Z, et al. Demographic characteristics and neuropsychological assessments of subjective cognitive decline (SCD) (plus). Ann Clin Transl Neurol 2020;7:1002–12. 10.1002/acn3.51068.

[52] Aghjayan SL, Buckley RF, Vannini P, Rentz DM, Jackson JD, Sperling RA, et al. The influence of demographic factors on subjective cognitive concerns and beta-amyloid. International Psychogeriatrics 2017;29:645–52. 10.1017/S1041610216001502.

[53] Lojo-Seoane C, Facal D, Guàrdia-Olmos J, Pereiro AX, Juncos-Rabadán O. Effects of Cognitive Reserve on Cognitive Performance in a Follow-Up Study in Older Adults With Subjective Cognitive Complaints. The Role of Working Memory. Front Aging Neurosci 2018;10:189. 10.3389/fnagi.2018.00189.

[54] Hong YJ, Park KW, Kang D-Y, Lee J-H. Prediction of Alzheimer’s Pathological Changes in Subjective Cognitive Decline Using the Self-report Questionnaire and Neuroimaging Biomarkers. Dementia and Neurocognitive Disorders 2019;18:19–29. 10.12779/dnd.2019.18.1.19.

[55] Bessi V, Mazzeo S, Padiglioni S, Piccini C, Nacmias B, Sorbi S, et al. From Subjective Cognitive Decline to Alzheimer’s Disease: The Predictive Role of Neuropsychological Assessment, Personality Traits, and Cognitive Reserve. A 7-Year Follow-Up Study. J Alzheimers Dis 2018. 10.3233/JAD-171180.

[56] Margolis SA, Kelly DA, Daiello LA, Davis J, Tremont G, Pillemer S, et al. Anticholinergic/Sedative Drug Burden and Subjective Cognitive Decline in Older Adults at Risk of Alzheimer’s Disease. The Journals of Gerontology: Series A 2020. 10.1093/gerona/glaa222.

[57] Molinuevo JL, Rabin LA, Amariglio R, Buckley R, Dubois B, Ellis KA, et al. Implementation of subjective cognitive decline criteria in research studies. Alzheimer’s & Dementia 2017;13:296–311. 10.1016/j.jalz.2016.09.012.

[58] Parfenov VA, Zakharov VV, Kabaeva AR, Vakhnina NV. Subjective cognitive decline as a predictor of future cognitive decline: a systematic review. Dement Neuropsychol 2020;14:248–57. 10.1590/1980-57642020dn14-030007.

[59] Slot RER, Sikkes SAM, Berkhof J, Brodaty H, Buckley R, Cavedo E, et al. Subjective cognitive decline and rates of incident Alzheimer’s disease and non-Alzheimer’s disease dementia. Alzheimers Dement 2019;15:465–76. 10.1016/j.jalz.2018.10.003.

[60] Beebe DW, Groesz L, Wells C, Nichols A, McGee K. The Neuropsychological Effects of Obstructive Sleep Apnea: A Meta-Analysis of Norm-Referenced and Case-Controlled Data. Sleep 2003;26:298–307. 10.1093/sleep/26.3.298.

[61] Health Quality Ontario. Vitamin B12 and cognitive function: an evidence-based analysis. Ont Health Technol Assess Ser 2013;13:1–45.

[62] McIntyre RS, Xiao HX, Syeda K, Vinberg M, Carvalho AF, Mansur RB, et al. The Prevalence, Measurement, and Treatment of the Cognitive Dimension/Domain in Major Depressive Disorder. CNS Drugs 2015;29:577–89. 10.1007/s40263-015-0263-x.

[63] Eckerström M, Göthlin M, Rolstad S, Hessen E, Eckerström C, Nordlund A, et al. Longitudinal evaluation of criteria for subjective cognitive decline and preclinical Alzheimer’s disease in a memory clinic sample. Alzheimers Dement (Amst) 2017;8:96–107. 10.1016/j.dadm.2017.04.006.

[64] Mazzeo S, Padiglioni S, Bagnoli S, Bracco L, Nacmias B, Sorbi S, et al. The dual role of cognitive reserve in subjective cognitive decline and mild cognitive impairment: a 7-year follow-up study. J Neurol 2019;266:487–97. 10.1007/s00415-018-9164-5.

[65] Tan X, Åkerstedt T, Lagerros YT, Åkerstedt AM, Bellocco R, Adami H-O, et al. Interactive association between insomnia symptoms and sleep duration for the risk of dementia—a prospective study in the Swedish National March Cohort. Age Ageing 2023;52:afad163. 10.1093/ageing/afad163.

[66] Ribaldi F, Rolandi E, Vaccaro R, Colombo M, Battista Frisoni G, Guaita A. The clinical heterogeneity of subjective cognitive decline: a data-driven approach on a population-based sample. Age Ageing 2022;51:afac209. 10.1093/ageing/afac209.

[67] Braak H, Braak E. Frequency of stages of Alzheimer-related lesions in different age categories. Neurobiol Aging 1997;18:351–7. 10.1016/s0197-4580(97)00056-0.

[68] Stewart R, Godin O, Crivello F, Maillard P, Mazoyer B, Tzourio C, et al. Longitudinal neuroimaging correlates of subjective memory impairment: 4-year prospective community study. Br J Psychiatry 2011;198:199–205. 10.1192/bjp.bp.110.078683.

