## Supplementary Materials for "Towards the development of a management protocol for Subjective Cognitive Decline: insights from a cross-sectional and longitudinal analyses of multimodal data from a memory clinic"

#### Supplementary Methods

##### 1. Neuropsychological assessment

The neuropsychological evaluation included: Mini-Mental State Examination (MMSE), tasks exploring verbal and spatial short-term and long-term verbal memory (five words and paired words acquisition and recall after 10 minute and after 24 hours; Rey Auditory Verbal Learning Test [1], Short Story Immediate and Delayed Recall [2], Rey-Osterrieth complex figure recall [3], Digit and Visuo-spatial Span forward and backward [4]), ecological memory (rivermead behavioral memory test (RBMT) [5]), attention (Trail Making Test A [6], attentional matrices [7], Multiple Features Targets Cancellation [8]), language (Category Fluency Task [9], Phonemic Fluency Task [1] and Italian language battery: Screening for Aphasia NeuroDegeneration [10]), constructional praxis (Copying drawings [1], Rey-Osterrieth complex figure copy [3], Clock test [11]) and executive function (Trail Making Test B [6], Stroop Test [12], Frontal Assessment Battery [13]).

**References**

[1] Carlesimo GA, Caltagirone C, Gainotti G. The Mental Deterioration Battery: normative data, diagnostic reliability and qualitative analyses of cognitive impairment. The Group for the Standardization of the Mental Deterioration Battery. Eur Neurol 1996;36:378–84. https://doi.org/10.1159/000117297.

[2] De Renzi E, Faglioni P, Ruggerini C. Prove di memoria verbale di impiego clinico per la diagnosi di amnesia 1977.

[3] Caffarra P, Vezzadini G, Dieci F, Zonato F, Venneri A. Rey-Osterrieth complex figure: normative values in an Italian population sample. Neurol Sci 2002;22:443–7. https://doi.org/10.1007/s100720200003.

[4] Monaco M, Costa A, Caltagirone C, Carlesimo GA. Forward and backward span for verbal and visuo-spatial data: standardization and normative data from an Italian adult population. Neurol Sci 2013;34:749–54. https://doi.org/10.1007/s10072-012-1130-x.

[5] Brazzelli M, Della Sala S, Laiacona M. Calibration of the Italian version of the Rivermead Behavioural Memory Test: A test for the ecological evaluation of memory. Bollettino Di Psicologia Applicata 1993:33–42.

[6] Giovagnoli AR, Del Pesce M, Mascheroni S, Simoncelli M, Laiacona M, Capitani E. Trail making test: normative values from 287 normal adult controls. Ital J Neurol Sci 1996;17:305–9. https://doi.org/10.1007/BF01997792.

[7] Della Sala S, Laiacona M, Spinnler H, Ubezio C. A cancellation test: its reliability in assessing attentional deficits in Alzheimer's disease. Psychol Med 1992;22:885–901. https://doi.org/10.1017/s0033291700038460.

[8] Marra C, Gainotti G, Scaricamazza E, Piccininni C, Ferraccioli M, Quaranta D. The Multiple Features Target Cancellation (MFTC): an attentional visual conjunction search

test. Normative values for the Italian population. Neurol Sci 2013;34:173–80. https://doi.org/10.1007/s10072-012-0975-3.
[9] Novelli G, Papagno C, Capitani E, Laiacona M. Tre test clinici di ricerca e produzione lessicale. Taratura su sogetti normali. / Three clinical tests to research and rate the lexical performance of normal subjects. Archivio Di Psicologia, Neurologia e Psichiatria 1970:477–506.
[10] Catricalà E, Gobbi E, Battista P, Miozzo A, Polito C, Boschi V, et al. SAND: a Screening for Aphasia in NeuroDegeneration. Development and normative data. Neurological Sciences : Official Journal of the Italian Neurological Society and of the Italian Society of Clinical Neurophysiology 2017;38:1469–83. https://doi.org/10.1007/s10072-017-3001-y.
[11] Shulman KI, Pushkar Gold D, Cohen CA, Zucchero CA. Clock-drawing and dementia in the community: A longitudinal study. International Journal of Geriatric Psychiatry 1993;8:487–96. https://doi.org/10.1002/gps.930080606.
[12] Caffarra P, Vezzadini G, Dieci F, Zonato F, Venneri A. Una versione abbreviata del test di Stroop: Dati normativi nella popolazione Italiana. Rivista di neurologia 2002;12:111–5.
[13] Appollonio I, Leone M, Isella V, Piamarta F, Consoli T, Villa ML, et al. The Frontal Assessment Battery (FAB): normative values in an Italian population sample. Neurol Sci 2005;26:108–16. https://doi.org/10.1007/s10072-005-0443-4.

#### Supplementary Tables

**Supplementary Table 1**. Regression coefficient (B) and significance level (p) per each variables influencing neuropsychological test scores.

| | Gender | Depressive symptoms | Insomnia | Anticholinergic use | BZD use |
|---|---|---|---|---|---|
| MMSE | | B = -0.08, p < 0.001 | | | |
| ROCFC | | B = -0.03, p < 0.001 | | | B = -0.24, p = 0.028 |
| TMT-B | | B = -0.01, p = 0.012 | B = -0.11, p = 0.039 | | |
| MCFT | | | | B = -0.70, p = 0.010 | B = -0.82, p = 0.003 |
| Spatial span | B = -0.30, p = 0.019 | | | | |
| Token test | B = -0.18, p = 0.006 | | | | |

**Supplementary Table 2**. Performance metrics for the best performing model. Values are reported as median [first percentile, third percentile] of the bootstrapped estimate (10e3 iterations)

| **Metric** | **Value** |
|---|---|
| Balanced accuracy | 74.37% [66.97, 80.90] |
| F1-score | 79.78% [ 72.61, 85.71] |
| Recall | 80.68% [71.59, 88.46] |
| Precision | 79.38% [69.89, 87.50] |
| AUC | 72.94% [63.77, 81.40] |

**Supplementary Table 3**. Concentrations of CSF biomarkers and AD biomarker positivity proportion

| | **Median (IQR)** | **min : max** |
|---|---|---|
| $A\beta_{42}$ (pg/mL) | 974.0 (654.0) | 435.0 : 1827.0 |
| $A\beta_{42}/A\beta_{40}$ | 0.09 (0.05) | 0.035 : 0.140 |
| p-tau (pg/mL) | 44.0 (25.4) | 14.0 : 124.0 |
| t-tau (pg/mL) | 338.0 (245) | 114 : 1048 |
| | **Frequencies** | **%** |
| Amyloid-PET | 9/23 | 39.1 |
| A+ | 14/43 | 32.6 |
| T+ | 9/37 | 24.3 |
| N+ | 11/37 | 29.7 |
| A+/T-/N- | 5/37 | 13.5 |
| A+/T+/N- | 2/37 | 5.4 |
| A+/T+/N+ | 6/37 | 16.2 |
| A-/T-/N+ | 3/37 | 8.1 |
| A-/T+/N+ | 1/37 | 2.7 |
| A-/T-/N- | 17/37 | 54.1 |

*The A+ proportion was calculated based on the 43 patients who underwent lumbar puncture and/or Amyloid-PET. The T+ and N+ proportions were calculated from patients who underwent CSF analysis.*

### Supplementary Figures

**Supplementary figure 1.** Comparison between weights of the edges in the network analysis. Black boxes indicate significant differences, gray boxes indicate non-significant differences

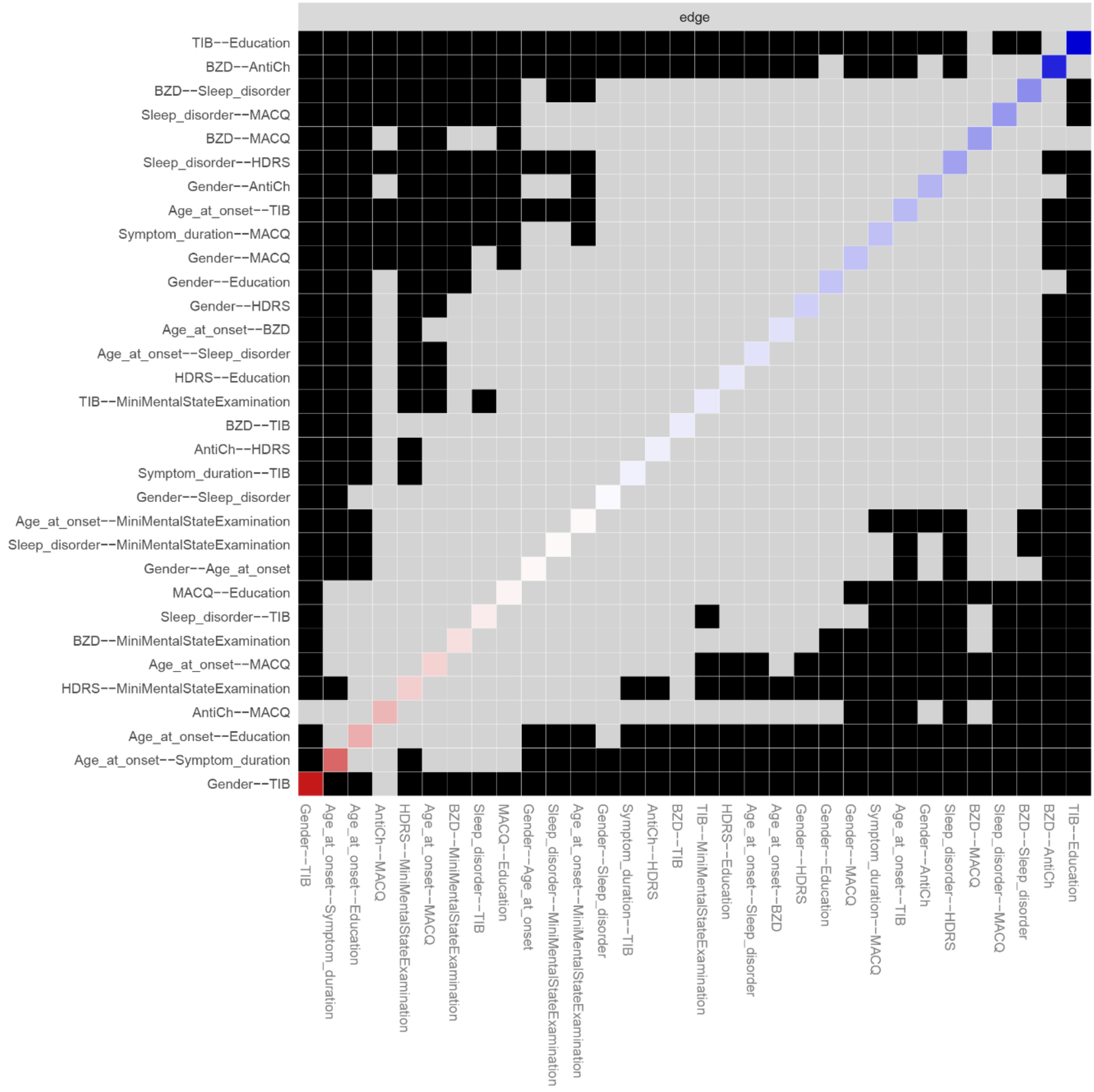

**Supplementary figure 2.** Cumulative incidence rate of progression to MCI per each year

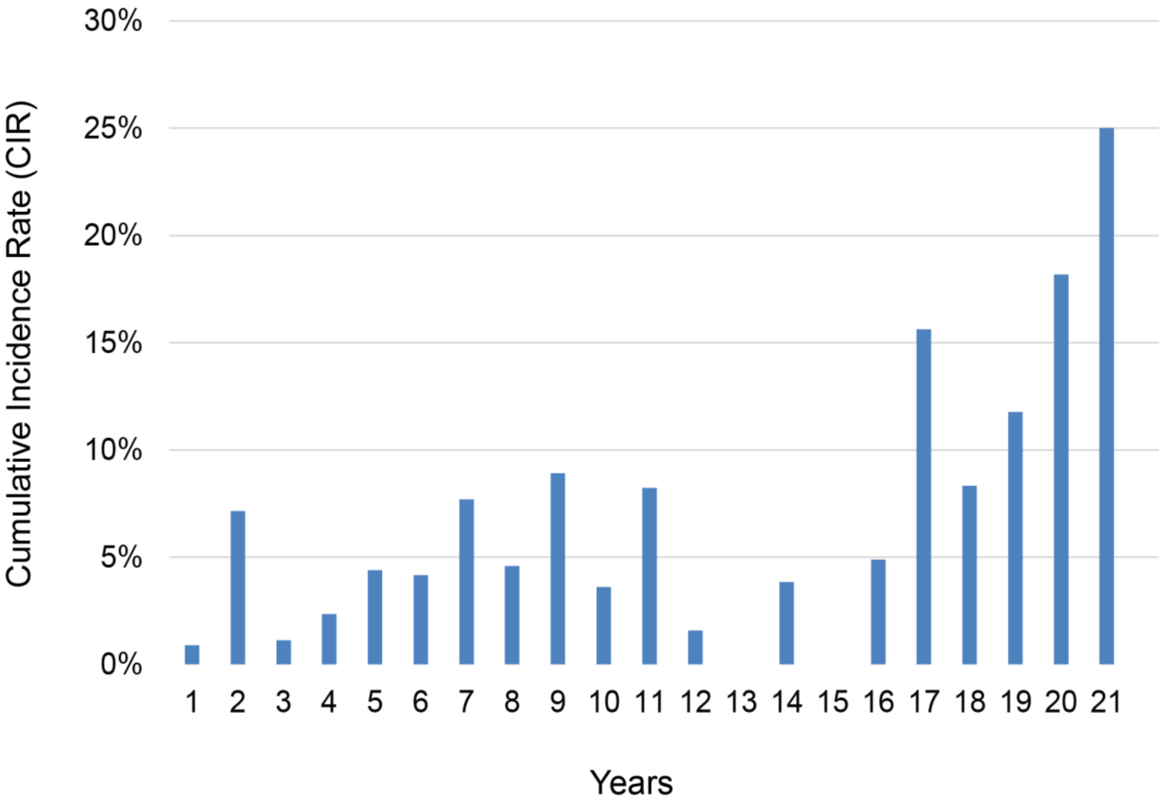

**Supplementary Figure 3**. Performance of the best performing model. (A) Confusion matrix. (B) ROC curve. Shaded area indicates 95% confidence interval, computed by bootstrapping (10e3 iterations). Green solid line indicates the mean of the bootstrapped ROC curve. Dashed grey line depicts the chance level line.

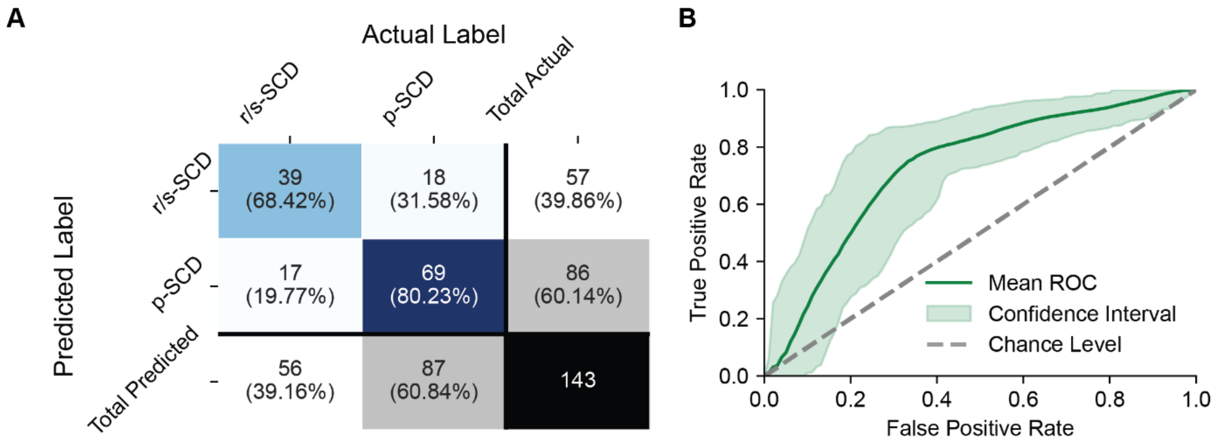

**Supplementary Figure 4**. Raw p-values of comparisons between pairs of features based on SHAP feature importance bootstrapping. Each square represents the binary significance of the comparison between two feature importance weights. The color map indicate the value of p-values.

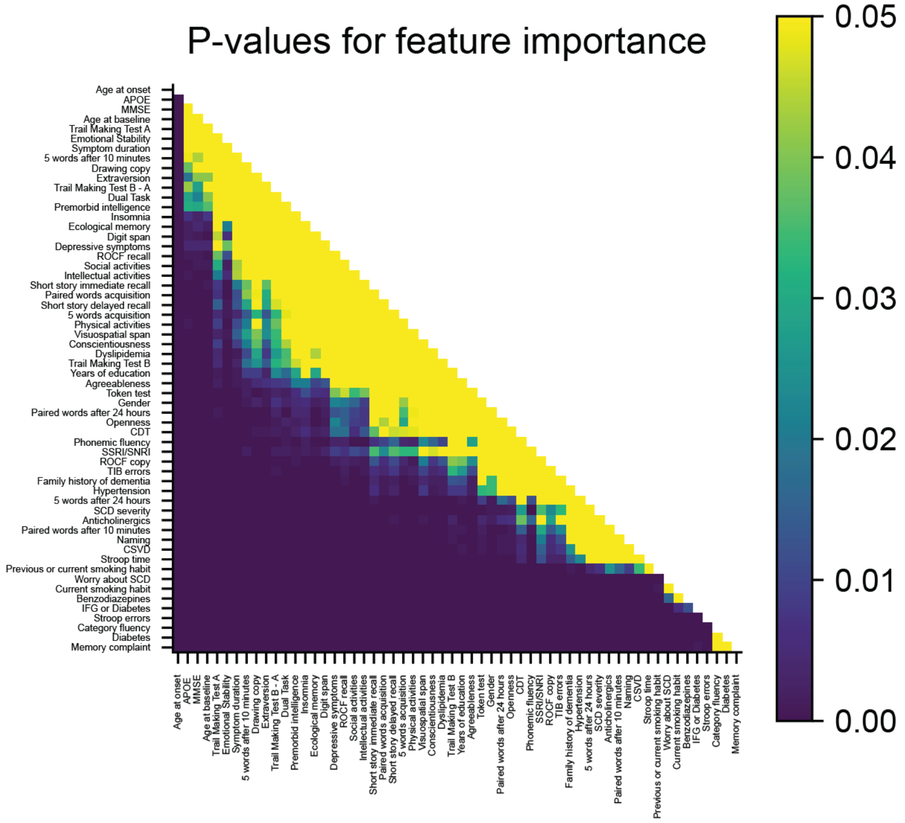

**Supplementary Figure 5**. Flowchart detailing the management protocol for SCD applied in our memory clinic.

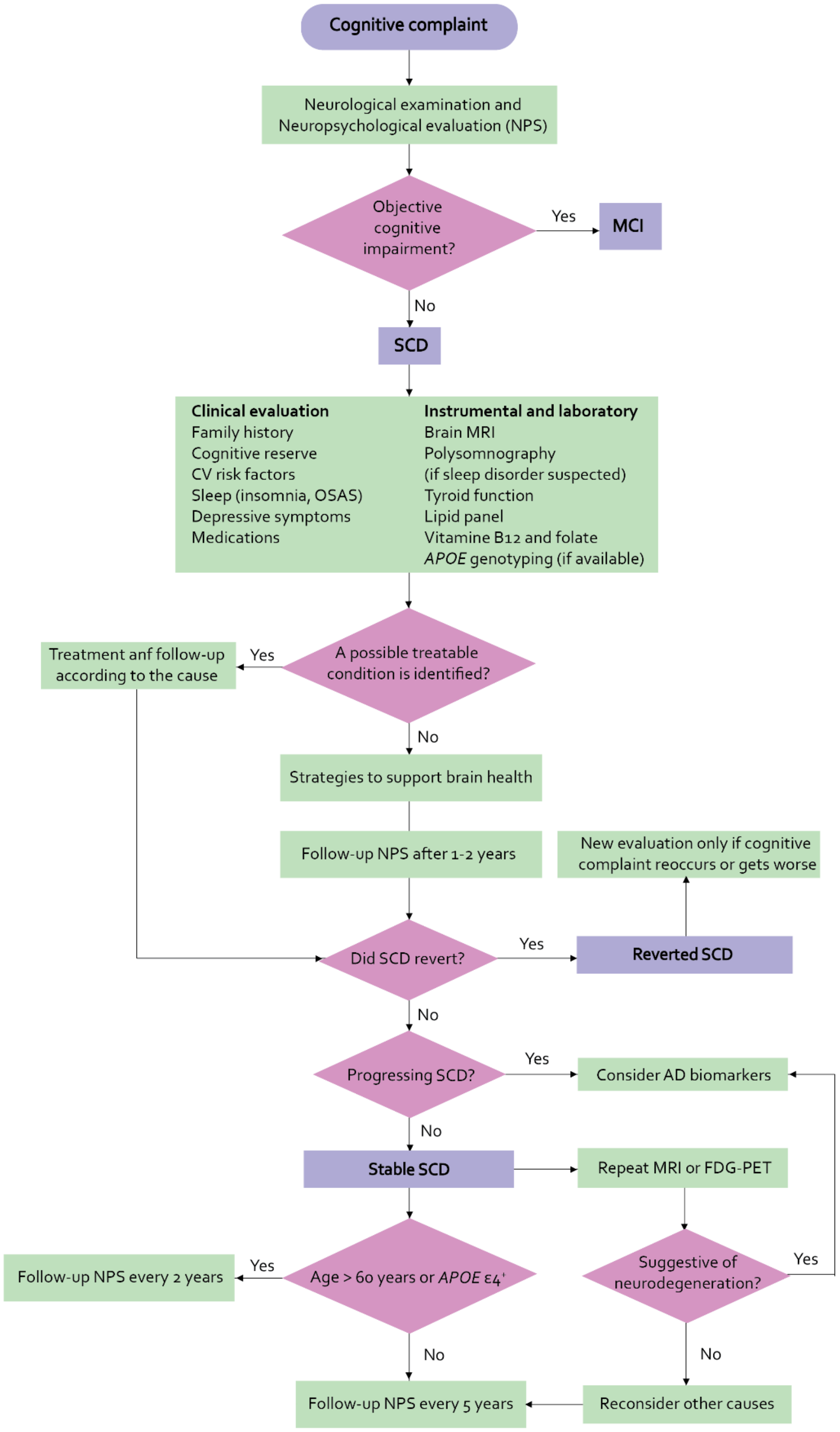
